# Preferences for benefit and harm outcomes of GLP-1 receptor agonists in adults with overweight or obesity: a multinational best–worst scaling study

**DOI:** 10.64898/2025.12.15.25342259

**Authors:** Hannah Moll, Milo A Puhan, Philipp Gerber, Felix Beuschlein, Katharina Frenes, Alessandra Spanu, Joan Walter, Henock G Yebyo

## Abstract

**Aims:** We aimed to elicit preferences for weight and harm outcomes of glucagon-like peptide-1 receptor agonist (GLP-1 RA) treatment. We examined heterogeneity across key patient subgroups to support tailored treatment decisions to identify patients most likely to benefit with minimal risks.

**Material and Methods:** We conducted a best–worst scaling survey of adults with overweight or obesity in 15 European countries, assessing 21 GLP-1 RA outcomesincluding 5% and 10% weight gain (to align with harms) and 19 adverse outcomes (eg. gastrointestinal events, hypoglycaemia, pancreatitis, gallbladder outcomes). Participants were provided lay descriptions of outcomes, rated seriousness on a visual analogue scale, and completed a best–worst scaling task. Preferences were estimated using a Bayesian hierarchical mixed multinomial logit model and rescaled from 0 (least worrisome) to 1 (most worrisome). Subgroup analyses were conducted based on sex, age, body mass index (BMI), and physical activity (36 subgroups).

**Results:** Among 2112 participants (48.1% female; mean age 39.51 years; median BMI 30.0 kg/m^2^), the least concerning outcomes were eructation (0.04) and flatulence (0.05), and the most concerning were pancreatitis (0.42), followed by 10% weight gain (0.41), cholecystitis (0.35), and cholelithiasis (0.32). Other outcome weights ranged from 0.11 to 0.25. Preference patterns were similar for extremes but varied by sex, age, BMI, and physical activity.

**Conclusions:** Preferences for GLP-1 RA outcomes varied across individuals and subgroups. Incorporating explicit patient preferences into routine care may better align GLP-1 RA prescribing with patient values and support individualised benefit–harm decisions.

## Introduction

The introduction of glucagon-like peptide-1 (GLP-1) receptor agonists (RA) and dual glucose-dependent insulinotropic polypeptide (GIP)/GLP-1 RA (collectively referred to as GLP-1 RAs) has shown significant promise in the treatment of obesity. While these drugs were originally approved for metabolic control in patients living with diabetes, they have shown substantial weight reduction in persons with obesity.^1–4^ In addition, recent studies have shown that these treatments can improve the prognosis of people living with cardiovascular disease, non-alcoholic fatty liver disease and sleep apnoea.^5–7^ However, GLP-1 RAs are also associated with a range of mild to moderate unwanted effects that challenge treatment compliance and effectiveness, including gastrointestinal events, pancreatitis, cholecystitis, as well as generalized adverse effects (harms), such as fatigue and headache, and hair loss.^8^ Multiple studies report that roughly 20 to 50% of patients on GLP-1 RAs discontinue treatment within the first year, most of which could be due to generalized malaise associated with adverse effects, high cost, or insufficient effectiveness.^9–11^ It is therefore important to assess patient preference trade-offs among GLP-1 RA-related positive and negative outcomes to guide tailored treatments.

A previous benefit-harm modelling study has shown a 10% weight loss to be net beneficial in treatment initiation, but the benefit-harm balance was highly sensitive to individual preferences, especially tolerance and risk levels for harm effects, as well as treatment goals. Although clinicians often discuss adverse effects with patients, such discussions are typically subjective, and weighing multiple benefits and harms alongside various patient characteristics is cognitively difficult. Systematic, quantitative incorporation of patient values into benefit–harm assessment and decision-making is needed. Quantifying the relative importance of outcomes—or integrating preference-elicitation tools into routine care—may help tailor treatment to patient profiles, improve adherence, and identify those most likely to benefit with acceptable risk.^12–14^ This study elicited preferences for weight (framed as 5% and 10% weight gain) and harm outcomes associated with GLP-1 RAs and examined subgroup heterogeneity in a multinational sample of adults living with overweight or obesity.

## Material and Methods

### Study design and participants

We conducted an online survey across multiple countries using best-worst scaling (BWS), a variant of a discrete choice experiment, among overweight and obese participants to elicit preferences for GLP-1 RA weight and harm outcomes across different subgroups. Eligible participants were 18 years or older, overweight or obese (body mass index (BMI) ≥ 25), and currently residing in Western or Northern Europe (Austria, Belgium, Denmark, France, Germany, Iceland, Ireland, Italy, Luxembourg, the Netherlands, Norway, Spain, Sweden, Switzerland or the UK).

There is no standard method for estimating sample size in BWS studies but the most frequently used sample size for overall sample size in health surveys were between 150 to 300. We targeted a sample of 2,160, with 60 respondents per predefined subgroup combination (sex × age × BMI × physical activity= 36 levels) to enable stable subgroup-specific preference estimates. To compensate for non-responses or incomplete questionnaires, we invited 2,500 participants to achieve the target sample size.

### Selection of outcomes

Based on our prior work, a review of the literature, and Swiss regulatory authority documents, we identified relevant benefit (weight reduction) and harm outcomes related to the GLP-1 RAs.^15^ While the benefit endpoint was weight reduction, we presented it as 5% and 10% weight gain so that all outcomes were expressed on a harm-comparable scale for consistent comparison. There are several harm outcomes reported in randomized controlled trials (RCTs) and other studies, but we narrowed the list down to the frequent (≥1%) and serious adverse effects as listed in the specialist information approved by the Swiss regulatory authority. Consequently, frequent harm outcomes included abdominal pain, constipation, diarrhoea, dizziness, dyspepsia, eructation, fatigue, flatulence, headache, hypoglycaemia, injection site reactions, nausea, upper respiratory tract infections, vomiting and moderate to serious harm outcomes included alopecia, cholecystitis, cholelithiasis and pancreatitis.

### Data collection

The survey collected socio-demographic data (age, sex, residence, education, employment status), clinical and lifestyle data (height, weight, physical activity, diet, medical conditions, medications), visual analogue scaling (VAS), and the BWS. First we provided clear, lay descriptions of each outcome to ensure consistent interpretation and measurement across individuals (see Supplementary Appendix). The lay descriptions focused on the signs and symptoms, treatment options, and prognosis. Participants completed both the VAS and BWS tasks after familiarization with these lay descriptions.

In the VAS taks, participants rated each outcome separately on a 0 (not concerning at all) to 100 scale (equivalent to having fatal or severe disabling stroke).^14^ These values were used to anchor relative preference scales from BWS to a natural scale of 0 to 1 (see analysis). Participants then completed a BWS (Case 1) questionnaire in which they selected the least and most concerning outcomes from a series of outcome combinations. Participants selected the least concerning and the most concerning outcome from each block (Supplementary Appendix). We generated the outcome combinations usng a balanced incomplete block design (BIBD)^15^, which ensures that each outcome appears the same number of times and co-appears with other outcomes in a balanced manner (see details in Supplementary Appendix).

The survey was developed in the Research Electronic Data Capture platform (REDCap) on a secure server. We pilot tested the questionnaire twice in English (n = 10 each) to ensure functionality and comprehensibility, then translated the final version into French, German, Italian and Spanish by native speakers. We recruited participants via Prolific® (https://www.prolific.com/), an online participant-recruiting platform, which allowed us to screen eligibility, monitor completion times, implement data quality controls, and return questionnaires for correction as needed.

### Data analysis

We converted the raw BWS data to a special matrix where we assigned 1 for outcomes selected as best, −1 for outcomes selected as worst, and 0 otherwise for all possible, pairwise combinations of responses. We used these data to estimate individual-level relative preference weights using Bayesian hierarchical mixed multinomial logit model, which produced preference estimates for each respondent.^16^ For interpretability and comparability, the resulting coefficients (on an arbitrary logit scale) were anchored to the VAS data to obtain preference weights on a 0 to 1 scale using hierarchical Bayesian linearmixed-effects model both at individual and subgroup level. This was preceded by correlation checks between the preference values and VAS scores using the Pearson correlation coefficient (Supplementary Appendix, Figure S1). The anchored weights were derived by inverse-logit-transforming the posterior predictions, providing anchored probabilities with corresponding 95% credible intervals.

Analyses were done for subgroups by age (18–29, 30–49, 50+), sex (male and female), BMI (25–29, 30–34, ≥35), physical activity of any type (<150 vs. ≥150 minutes per week). For each predefined subgroup, we computed Bayesian coefficients of determination (R^2^) from the hierarchical anchoring model using posterior predictions restricted to that subgroup (Supplementary Appenidx, Table S2).

To assess whether participants answered questions consistently, we duplicated BWS questions randomly without indicating this for the respondents. This allowed us to compare if participants selected the same most concerning and least concerning outcomes in both instances. For each repeated pair, we cross-tabulated responses and calculated the proportion of consistent responses.

All analyses were performed in R (version 4.5.1).

### Ethics

Ethical approval for this study was waived by the Cantonal Ethics Committee of Zurich, Req-2025-01495.

### Role of funding

The funder of this study had no role in study design, data collection, data analysis, data interpretation, writing of the report and the decision to submit the report for publication.

## Results

### Sample characteristics

We obtained responses from 2112 participants (98% of the target sample), based on eligibility and complete questionnaire responses. The sex distribution was almost equal with 1015 female participants (48.1%), a mean age of 39.51 (SD 13.05) and a median BMI of 30.0 (27.4 to 34.0). Most of the participants were educated, with 854 (40.4%) having completed a secondary education and 1218 (57.7%) having completed a university degree and most were employed (1529, 72.4%). A little less than half of the participants reported to be physically active for more than 150 minutes per week (951, 45.0%) while only 695 (32.9%) reported following a diet. Approximately half the participants (1021, 48.3%) took at least one medication per day. The most common places of residence were the UK (893, 42.3%), Germany (336, 15.9%) and Italy (218, 10.3%).

### Preference results

Preference results for the overall sample from the VAS, BWS, and anchored weights are shown in Table 2. The mean VAS of the outcomes ranged from 0.10 to 0.46 on a 0 to 1 scale. Participants ranked eructation (0.10, 0.09 to 0.10), flatulence (0.10, 0.09 to 0.11) and constipation (0.17, 0.16 to 0.18) as least concerning and weight gain of 10% (0.46, 0.45 to 0.47), pancreatitis (0.40, 0.39 to 0.41), cholecystitis (0.35, 0.34 to 0.36), and cholelithiasis (0.33, 0.32 to 0.34) as most concerning outcomes. The preference coefficients from the BWS data using hierarchical Bayesian conditional logit model were consistent with the VAS ranking (Table 2) and were strongly correlated with the VAS scores (r = 0.81; see Supplementary Appendix, Figure S1). The anchored weights derived from preference coefficients and VAS ranged from eructation (preference weight 0.04; 95% CI 0.02 to 0.07) and flatulence (0.05; 95% CI 0.03 to 0.08) to pancreatitis (0.42; 95% CI 0.34 to 0.51), 10% weight gain (0.41; 95% CI, 0.34 to 0.52), cholecystitis (0.35; 95% CI 0.28 to 0.42), and cholelithiasis (0.32; 95% CI, 0.25 to 0.40).

**Table 1.**
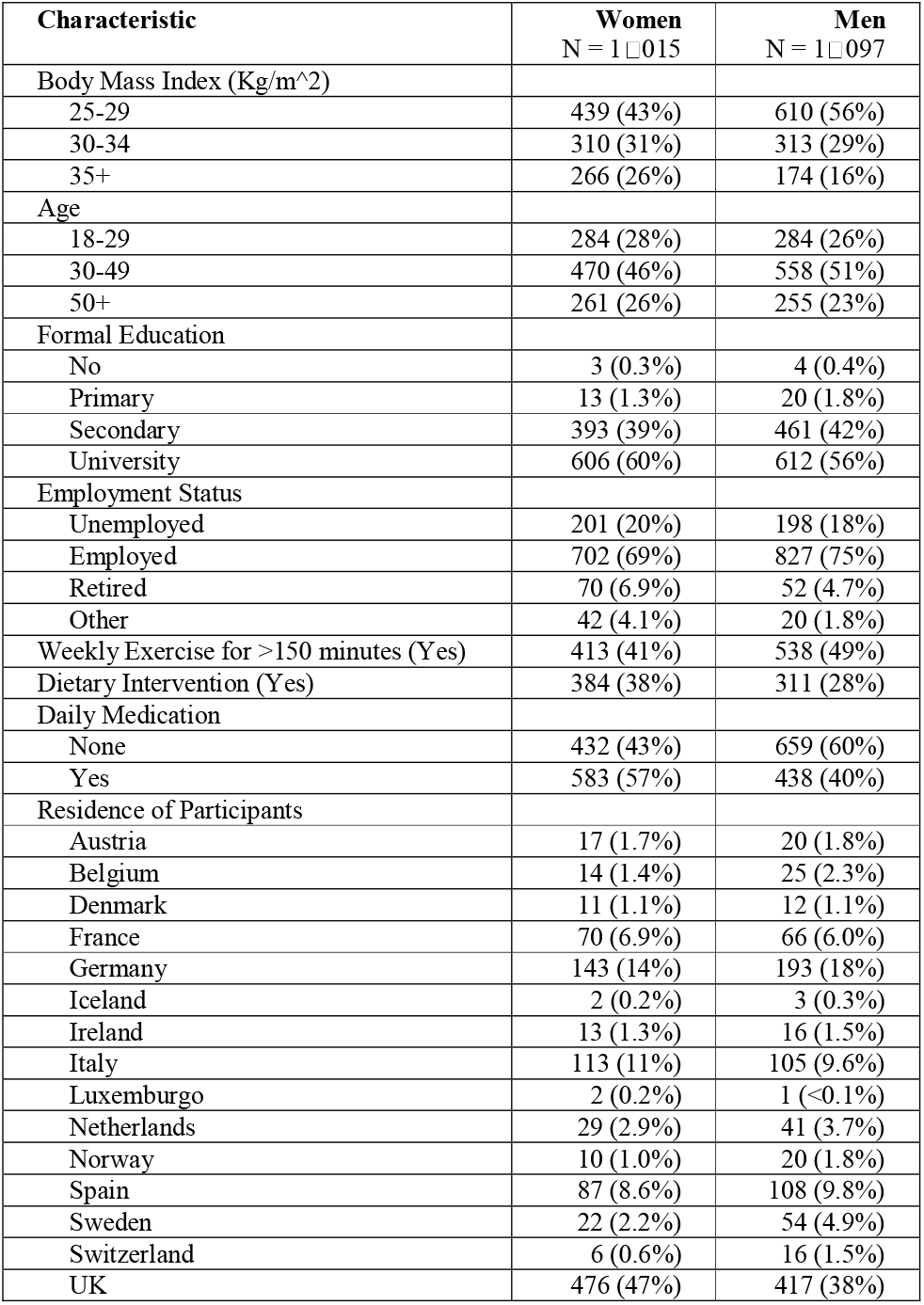
Characteristics of respondents.

**Table 2:** Preference estimates of the overall sample: Mean visual analogue scales, conditional logit coefficients, and anchored preference weights.

| Outcomes | Mean VAS (95% CI) | Preference coefficient (95% CI) | Anchored weight (95% CI) |
| --- | --- | --- | --- |
| Eructation | 0.10 (0.09 to 0.11) | -5.72 (-5.76 to -5.68) | 0.04 (0.02 to 0.07) |
| Flatulence | 0.10 (0.09 to 0.11) | -4.91 (-4.94 to -4.87) | 0.05 (0.03 to 0.08) |
| Constipation | 0.17 (0.16 to 0.18) | -2.09 (-2.13 to -2.06) | 0.11 (0.08 to 0.16) |
| Injection site reactions | 0.16 (0.15 to 0.17) | -1.83 (-2.17 to -1.49) | 0.11 (0.06 to 0.15) |
| Indigestion | 0.19 (0.18 to 0.20) | -1.98 (-2.08 to -1.88) | 0.12 (0.09 to 0.17) |
| Fatigue | 0.22 (0.21 to 0.23) | -1.76 (-1.83 to -1.70) | 0.13 (0.10 to 0.19) |
| Headache | 0.22 (0.21 to 0.23) | -2.14 (-2.21 to -2.08) | 0.13 (0.09 to 0.18) |
| Nausea | 0.18 (0.17 to 0.19) | -1.45 (-1.54 to -1.36) | 0.13 (0.09 to 0.17) |
| Alopecia | 0.22 (0.21 to 0.23) | -0.91 (-1.30 to -0.52) | 0.14 (0.09 to 0.19) |
| Diarrhea | 0.19 (0.18 to 0.20) | -1.04 (-1.08 to -0.99) | 0.14 (0.10 to 0.19) |
| Vomiting | 0.24 (0.23 to 0.25) | 0.50 (0.46 to 0.53) | 0.19 (0.14 to 0.25) |
| Abdominal pain | 0.23 (0.22 to 0.24) | 1.23 (1.09 to 1.36) | 0.20 (0.15 to 0.26) |
| Hypoglycemia | 0.26 (0.25 to 0.27) | 0.33 (0.29 to 0.36) | 0.20 (0.14 to 0.26) |
| Dizziness | 0.27 (0.26 to 0.28) | 0.21 (0.04 to 0.39) | 0.21 (0.17 to 0.28) |
| Upper respiratory tract infection | 0.23 (0.22 to 0.24) | 1.98 (1.92 to 2.04) | 0.23 (0.14 to 0.29) |
| Weight gain of 5% | 0.31 (0.30 to 0.32) | 0.87 (0.71 to 1.03) | 0.24 (0.20 to 0.31) |
| Upper abdominal pain | 0.27 (0.26 to 0.28) | 2.06 (2.01 to 2.11) | 0.25 (0.19 to 0.30) |
| Cholelithiasis | 0.33 (0.32 to 0.34) | 3.43 (3.35 to 3.50) | 0.32 (0.25 to 0.40) |
| Cholecystitis | 0.35 (0.34 to 0.36) | 3.93 (3.87 to 4.00) | 0.35 (0.28 to 0.42) |
| Weight gain of 10% | 0.46 (0.45 to 0.47) | 3.80 (3.73 to 3.88) | 0.41 (0.34 to 0.52) |
| Pancreatitis | 0.40 (0.39 to 0.41) | 5.49 (5.37 to 5.61) | 0.42 (0.34 to 0.51) |

VAS: Visual analogue scale

### Subgroup differences

Figure 1A shows the overall range of preference weights across all subgroups for each outcome. While heterogeneity was modestly reduced within subgroups defined by sex, age, BMI, and physical activity compared with individual-level preferences (Supplementary Appendix Figure S2), substantial variability remained for several outcomes. Figure 1B displays the distribution of subgroup-specfic preference weights stratified by sex, age and BMI for the two benefit outcomes and the four harm outcomes exhibiting the greatest heterogeneity in preferences across subgroups. Physical activity was not included in Figure 1B because weights differed minimally by physical activity. Detailed subgroup specific estimates are provided in Table S1 in the Supplementary Appendix.

**Figure 1:**
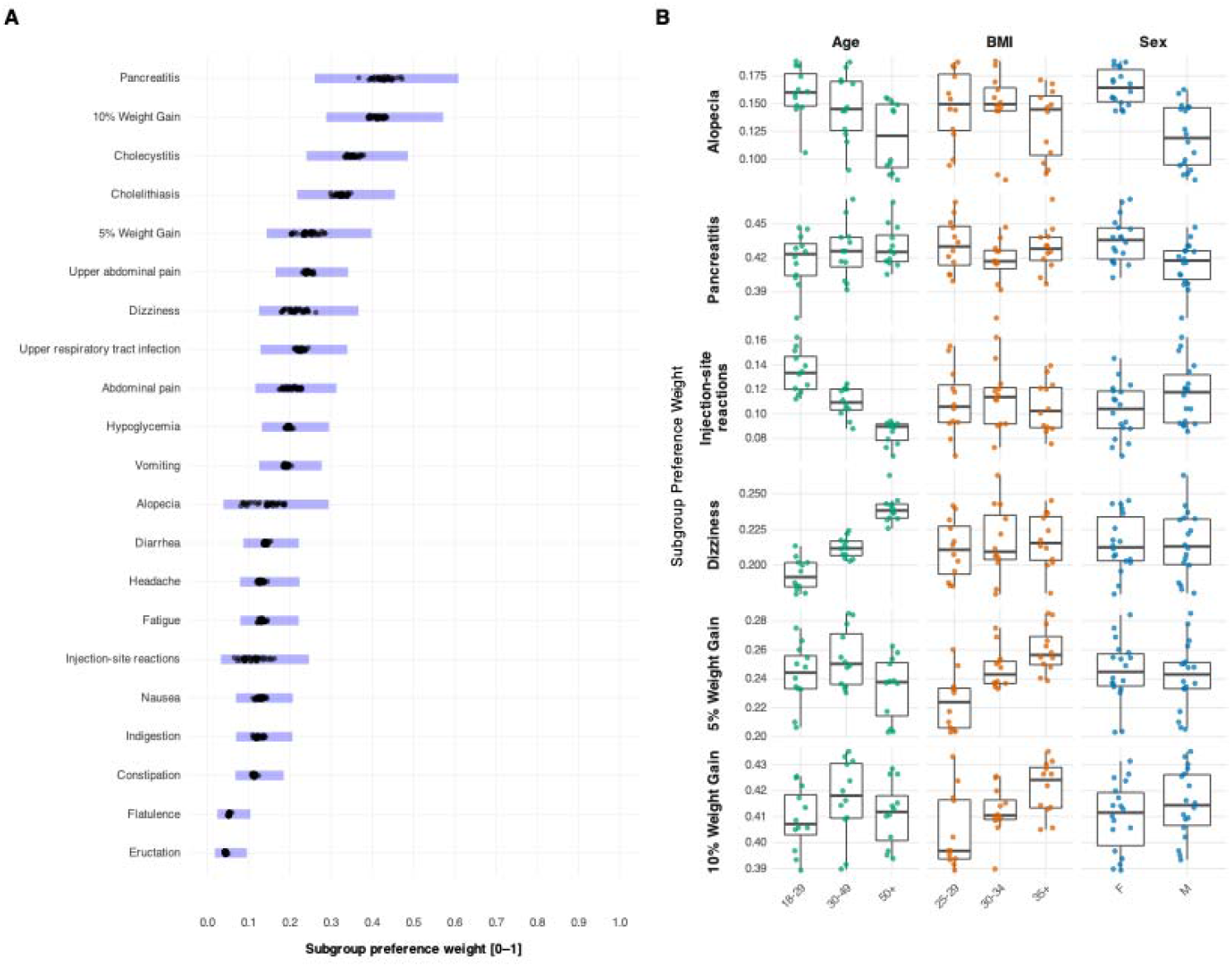
Subgroup preferences for GLP-1 RA outcomes. (A) Subgroup-aggregated preference weights. Points represent subgroup estimates for the 36 prespecified subgroups (sex × age × BMI × physical activity). Bands indicate the range of lower and upper 95% credible-interval across subgroups for each outcome (0 = not concerning; 1 = most concerning). (B) Distribution of subgroup-specific preference weights stratified by sex, age group, and BMI for the two benefit outcomes and the four harm outcomes with the greatest between-subgroup heterogeneity in preference weights. Physical activity is not included because differences in preference weights by physical activity level were minimal.

While the least and most worrisome outcomes remained generally consistent across subgroups, the magnitudes of the associated preference weight varied considerably. Across the 36 subgroup levels, variability was modest for milder gastrointestinal symptoms—e.g., eructation (0.04–0.05), flatulence (0.05–0.06), and constipation (0.11–0.12)—and larger for relatively severe outcomes—e.g., 10% weight gain (0.39–0.43), pancreatitis (0.37– 0.47), cholecystitis (0.33–0.38), and alopecia (0.08–0.19). Overall, younger participants tended to be more concerned about outcomes requiring more intensive treatment, such as cholecystitis and cholelithiasis, as well as outcomes such as alopecia and injection-site reactions. For instance, younger women expressed greater concern about alopecia, whereas men, particularly older men, were comparatively less concerned. In addition, older participants were more concerned about outcomes affecting daily functioning, including dizziness, headache, and fatigue. Individuals with a BMI above 35 rated a 10% weight as more concerning compared with individuals with a lower BMI (see details in Supplementary Appendix, Table S1). While the sample size was sufficient in most subgroups, some of them had only a few participants that led to wider preference estimates.

The hierarchical model showed a moderate to good overall fit, with Bayesian R^2^ ranging from 0.32 to 0.78 across subgroups (median 0.61; see Supplementary Appendix Table S2).

### Consistency

To evaluate response consistency in the BWS questionnaire, we compared answers to two repeated questions. Figure 2 presents response patterns with diagonal cells indicating consistent responses between the original and repeated questions both for the most concerning and least concerning outcomes. The high proportion (in blue) showed strong concistency between the repeated questions. Moderate inconsistencies were observed for a few outcomes, such as injection site reactions and eructation in the most concerning choices as well as weight gain, pancreatitis and cholelithiasis in the least concerning choices. These inconsistencies may reflect response errors.

**Figure 2:**
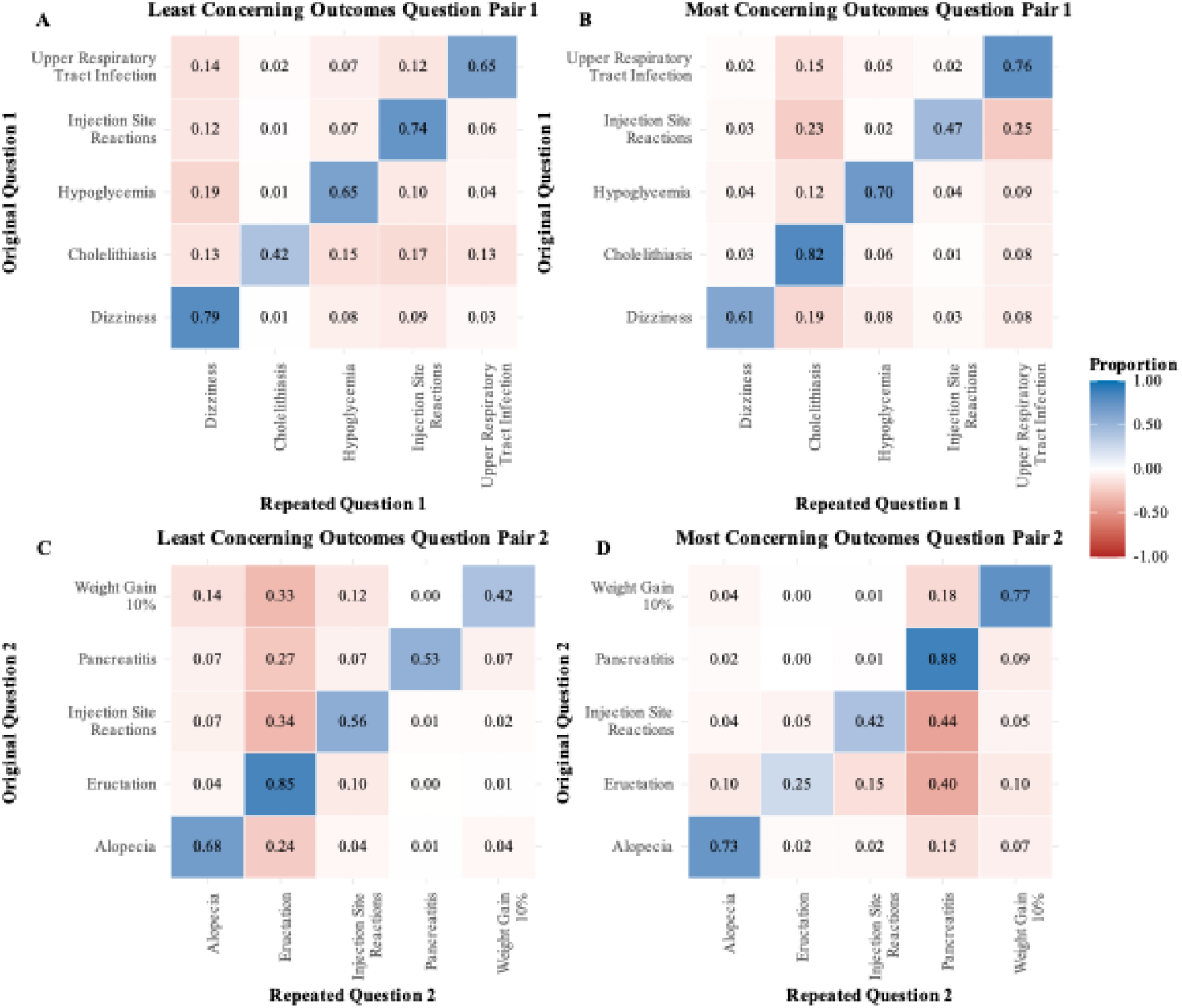
Consistency of responses. Display of proportion of consistent responses across repeated question pairs. Diagonal blue cells indicate consistent responses, and off-diagonal red cells denote inconsistencies; darker shading corresponds to higher proportions. Consistency was assessed separately for least concerning (A, C) and most concerning (B, D) outcomes.

## Discussion

This study provides empirical preference weights for GLP-1 RA-related outcomes in a multinational population of adults living with overweight or obesity. Our results showed considerable heterogeneity in these preferences across participant subgroups. Although most gastrointestinal□related harms had preference weights in the mild□to□moderate range, a 10% weight gain, pancreatitis and cholecystitis fell at the upper end, reflecting their greater perceived seriousness. These outcomes were likely received as more concerning to participants, as they could substantially impact health-related quality of life and, in rare cases, may even require surgical intervention. However, milder outcomes, such as eructation, nausea, vomiting, and diarrhoea, could be equally important in decision making, since they occur more frequently, easily disrupt daily functioning, and collectively interfere with quality of life.

Different studies in various areas show that individual preferences generally exhibit high heterogeneity and are not reliably predicted only by sociodemographic or clinical characteristics.^17,18^ Generally, observable variables often explain only a small fraction of between-person variation. Concistent with this literature, we observed similar challenges in this study wide dispersion in preferences for most outcomes across individuals, even after stratification by age, sex, BMI, and physical activity. These factors explained some of the heterogeneities, but left most variation unexplained. For example, a 10% weight gain elicited the most concern among younger individuals with a BMI over 35, possibly reflecting the greater personal and social relevance of weight-related outcomes for this subgroup. Women expressed greater concern about alopecia than men, which may reflect cosmetic considerations, particularly among younger individuals. Variation across subgroups was evident for most outcomes and substantial across individuals. These findings imply the need to incorporate patient preferences into clinical decision-making to improve adherence, align decisions with patient values, and help to optimize resources.

Comparisons of our results with existing studies are limited by sparse evidence and by difference in decision contexts. The Global Burden of Disease (GBD) study, for example, conducted large-scale international surveys to derive disability weights for use in calculating global disability-adjusted life years.^19^ Although our preference weights broadly align with the corresponding GBD disability weights, direct one-to-one comparison was not feasible given the different aims, designs, contexts, and populations. To our knowledge, this is the first large-scale preference elicitation survey for GLP-1 RA treatments. They can also be used in quantitative benefit–risk assessments of GLP-1 RAs, in which evidence on benefits and harms is weighted by the estimated preference weights to estimate overall treatment impact and identify patients or subgroups with the greatest expected net benefit, as shown in different studies^12–14,20–22^.

In our earlier benefit–harm modeling,^13^ for instance, generic (non-empirical) outcome weights were used due to lack of preference data, and the resulting benefit–harm balance was highly sensitive to the preference weights— underscoring the importance of aligning decisions with the preferences of individuals or clinically relevant subgroups to achieve optimal outcomes. Our findings may help explain why GLP-1 RAs exhibit high discontinuation rates despite documented weight-loss efficacy.^10,11,23,24^ Studies show about 65% of GLP-1 RA users discontinue therapy within the first year, with even greater attrition thereafter.^24^ While multiple factors— such as financial constraints or lack of desired weight loss—contribute to dropout, a substantial share is attributable to general discomfort from adverse effects.^24^ Accordingly, it is essential to develop tools that personalize treatment choice by aligning patients’ therapeutic goals and their individual benefit-harm profiles.

Although our study provides valuable insights into subgroup preferences from a relatively large cohort, some limitations warrant consideration. Individuals with lower educational attainment were underrepresented on the Prolific platform, as is commonly the case in research, and their outcome preferences may differ from those of higher-educated participants. Moreover, nearly half of the participants were currently residing in the UK, which could introduce a limitation, as healthcare systems, cultural norms, and clinical priorities may vary across countries. However, recent studies have found limited variation in preferences across different social and geographic settings.^14,25^ Further investigation—including qualitative research—is needed to elucidate the sources of the remaining heterogeneity and to assess the extent to which preferences are truly individual. Studies have to also include examining the impact of outcome preference variation in treatment outcomes by incorporating them to decision-making. The University of Zurich and the University Hospital of Zurich have developed a decision aid (currently under pilot testing) based on a benefit-harm blanace model that takes into account patient risks, GLP-1 RA evidence on benefit and harm outcomes, and preferences to support decision-making for weight management^26^.

In conclusion, this preference elicitation study demonstrated considerable variability in how individuals and subgroups value treatment outcomes commonly associated with GLP-1 receptor agonists. While a portion of this heterogeneity can be explained by subgroup characteristics, some variability remains unexplained, warranting further research into how subgroup-specific patterns might be used to support or complement individual-level assessments. The findings underscore the importance of incorporating patient preferences into clinical decision-making for tailored treatments, potentially improving treatment adherence, and minimizing avoidable harms and costs.

## Data Availability

De-identified participant data will be made available upon reasonable request to bona fide researchers who submit a research proposal for ethically approved purposes. Access will require a data-sharing agreement, and requests will be reviewed on a case-by-case basis.

## Contributors

HM, HGY and MAP conceived and designed the study. HM and HGY developed the survey instrument and implemented it using REDCap. HM and HGY aquired and analysed the data and prepared the manuscript. HM, MAP, HGY, PG, FB, KF, AS and JW critically revised the manuscript for important intellecutal content and approved the final manuscript. HM and HGY collected, analysed and verified the data. HGY and HM attest that all listed authors meet authorship criteria and that no others meeting the criteria have been omitted. All authors confirm the final draft and its submission.

## Acknowledgements

This study was financially supported by grants from The LOOP Zurich (FB and MAP).

