## Supplementary Appendix for "Preferences for benefit and harm outcomes of GLP-1 receptor agonists in adults with overweight or obesity: a multinational best–worst scaling study"

#### Preference elicitation questionnaire

##### Part I: Socio-demographic data

LOOBESITY ZURICH | Universität Zürich | USZ Universitäts Spital Zürich | THE LOOP ZURICH

A A A
English

**Welcome!**

Thank you for taking part in this survey. In this first part, we'll ask a few simple questions about you. This helps us understand more about the people taking part in our research.

Once you finish, you'll automatically move to the next part of the survey. **If you are completing the survey on a mobile phone, you may need to rotate your device to ensure some questions are formatted correctly.** You can change the survey language in the top right corner of the page.

Please answer honestly.

Thank you for your time!

**Part 1 of 3**

**Please enter your unique Prolific ID**
* must provide value

**Please enter your age**
* must provide value

**Please select your biological sex**
* must provide value

- ○ Male
- ○ Female
- ○ Other

reset

**Where do you currently live?**
* must provide value

**What is your native language?**
* must provide value

**How tall are you (in m)?**
E.g. 1.60
* must provide value

**What is your weight (in kg)?**
E.g. 70
* must provide value

**Do you have any of the following medical conditions? Please check all that apply:**
* must provide value

- ☐ Cardiovascular disease (e.g., heart disease, stroke)
- ☐ Chronic kidney disease
- ☐ Dyslipidemia
- ☐ High cholesterol
- ☐ High blood pressure
- ☐ Non-alcoholic fatty liver disease
- ☐ Osteoarthritis or joint issues
- ☐ Polycystic ovary syndrome (PCOS)
- ☐ Sleep apnea or other breathing disorders
- ☐ Other
- ☐ None

**What is your current employment status?**
* must provide value

- ○ Unemployed
- ○ Employed
- ○ Employed part-time
- ○ Employed with a temporary contract
- ○ Self-employed
- ○ Retired
- ○ Student
- ○ Other

reset

**What is the highest level of education you have completed?**
* must provide value

- ○ No formal education completed
- ○ Primary Education (e.g. Primary School)
- ○ Secondary Education (e.g. High School, Vocational Education)
- ○ University Degree

reset

**How much time do you usually spend on physical activity per week?**
**E.g. Biking, Swimming, Running**
* must provide value

- ○ Less than 150 minutes per week
- ○ Between 150 to 300 minutes per week
- ○ More than 300 minutes per week

reset

**Do you have any disabilities that impact your daily life?**
* must provide value

- ○ Yes
- ○ No
- ○ Prefer not to say

reset

**Are you currently following a diet?**
* must provide value

- ○ Yes
- ○ No

reset

**How many medications do you take daily?**
* must provide value

- ○ None
- ○ 1-2
- ○ 3-5
- ○ 6 or more

reset

**Have you undergone any past treatments for obesity? (Select all that apply)**
* must provide value

- ☐ No, I have not received any treatments
- ☐ Diet and lifestyle changes
- ☐ Prescription medications for weight loss (Saxenda, Wegovy, Mounjaro)
- ☐ Bariatric surgery (e.g., gastric bypass, sleeve gastrectomy)
- ☐ Other

Continue

#### Part II: Lay descriptions and visual analogue scaling

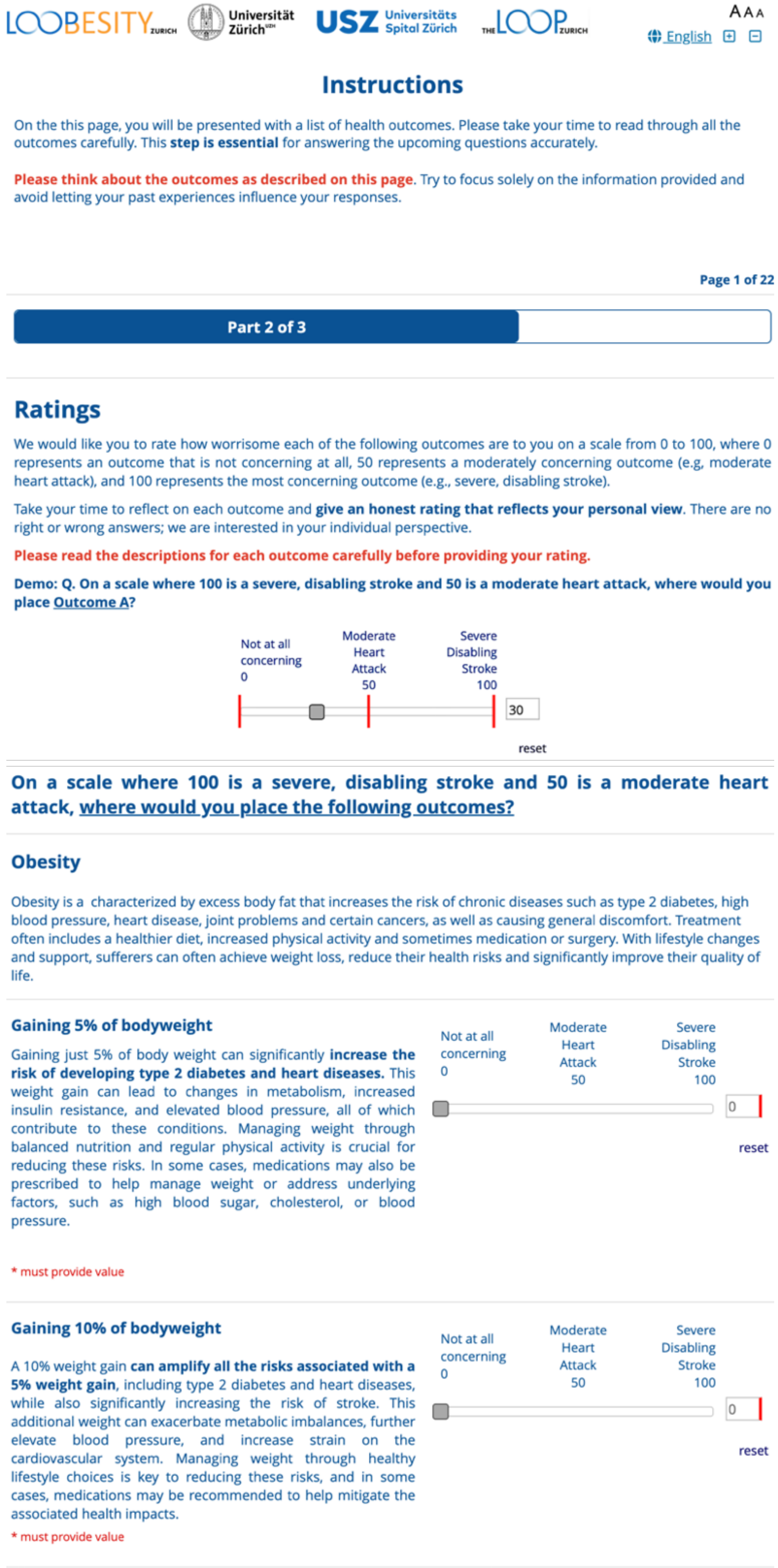

AAA

English

### Instructions

On the this page, you will be presented with a list of health outcomes. Please take your time to read through all the outcomes carefully. This **step is essential** for answering the upcoming questions accurately.

**Please think about the outcomes as described on this page**. Try to focus solely on the information provided and avoid letting your past experiences influence your responses.

**Page 1 of 22**

**Part 2 of 3**

#### Ratings

We would like you to rate how worrisome each of the following outcomes are to you on a scale from 0 to 100, where 0 represents an outcome that is not concerning at all, 50 represents a moderately concerning outcome (e.g, moderate heart attack), and 100 represents the most concerning outcome (e.g., severe, disabling stroke).

Take your time to reflect on each outcome and **give an honest rating that reflects your personal view**. There are no right or wrong answers; we are interested in your individual perspective.

**Please read the descriptions for each outcome carefully before providing your rating.**

**Demo: Q. On a scale where 100 is a severe, disabling stroke and 50 is a moderate heart attack, where would you place Outcome A?**

Not at all concerning 0 | Moderate Heart Attack 50 | Severe Disabling Stroke 100

30

reset

##### On a scale where 100 is a severe, disabling stroke and 50 is a moderate heart attack, where would you place the following outcomes?

##### Obesity

Obesity is a characterized by excess body fat that increases the risk of chronic diseases such as type 2 diabetes, high blood pressure, heart disease, joint problems and certain cancers, as well as causing general discomfort. Treatment often includes a healthier diet, increased physical activity and sometimes medication or surgery. With lifestyle changes and support, sufferers can often achieve weight loss, reduce their health risks and significantly improve their quality of life.

**Gaining 5% of bodyweight**

Gaining just 5% of body weight can significantly **increase the risk of developing type 2 diabetes and heart diseases.** This weight gain can lead to changes in metabolism, increased insulin resistance, and elevated blood pressure, all of which contribute to these conditions. Managing weight through balanced nutrition and regular physical activity is crucial for reducing these risks. In some cases, medications may also be prescribed to help manage weight or address underlying factors, such as high blood sugar, cholesterol, or blood pressure.

Not at all concerning 0 | Moderate Heart Attack 50 | Severe Disabling Stroke 100

0

reset

* must provide value

**Gaining 10% of bodyweight**

A 10% weight gain **can amplify all the risks associated with a 5% weight gain**, including type 2 diabetes and heart diseases, while also significantly increasing the risk of stroke. This additional weight can exacerbate metabolic imbalances, further elevate blood pressure, and increase strain on the cardiovascular system. Managing weight through healthy lifestyle choices is key to reducing these risks, and in some cases, medications may be recommended to help mitigate the associated health impacts.

Not at all concerning 0 | Moderate Heart Attack 50 | Severe Disabling Stroke 100

0

reset

* must provide value

#### Gallbladder Inflammation (Cholecystitis)

Gallbladder inflammation is **swelling or irritation of the gallbladder that can result from gallstones, digestive fluids, or medications**. Common symptoms include right upper abdominal pain or pain in the right shoulder, nausea, vomiting, and fever. It is usually mild to moderate in severity and may require treatments such as intravenous fluids, antibiotics, pain relievers, or the removal of gallstones if they are the cause. If caused by medications, discontinuing the medication may help, but this should be carefully considered if the medication is essential for other purposes.

* must provide value

| Not at all concerning 0 | Moderate Heart Attack 50 | Severe Disabling Stroke 100 |
|---|---|---|

0

reset

<< Previous Page Next Page >>

#### Inflammation of the Pancreas (Pancreatitis)

Pancreatitis is the **inflammation or irritation of the pancreas**. Common symptoms include abdominal pain that may radiate to the back, nausea, vomiting, and fever, typically presenting as mild to moderate in severity. Causes may include medications, gallstones, heavy alcohol use, or other factors. Management often involves dietary improvements, avoiding alcohol and smoking, discontinuing triggering medications if possible, and, in some cases, administering intravenous fluids and pain relievers.

* must provide value

| Not at all concerning 0 | Moderate Heart Attack 50 | Severe Disabling Stroke 100 |
|---|---|---|

0

reset

<< Previous Page Next Page >>

#### Abdominal Pain

Abdominal pain refers to any **localized or diffused discomfort or pain in your belly**. It can be caused by medications, problems with the digestive tract from the chest to the pelvis, or issues with the abdominal muscles. Abdominal pain often presents as mild but can sometimes be moderate. It can be alleviated with some over-the-counter pain medications or home remedies or self-care measures, such as adjusting diet and staying hydrated.

* must provide value

| Not at all concerning 0 | Moderate Heart Attack 50 | Severe Disabling Stroke 100 |
|---|---|---|

0

reset

<< Previous Page Next Page >>

#### Headache

A headache is **pain in the head or face**, often mild to moderate in severity and caused by various factors, including medications. Mild headaches can often be alleviated with non-pharmacological remedies, such as applying heat or cold packs, neck massage, or resting. Over-the-counter painkillers may be necessary if the headache disrupts daily activities. If the cause is known, such as medications, discontinuing them might help.

* must provide value

| Not at all concerning 0 | Moderate Heart Attack 50 | Severe Disabling Stroke 100 |
|---|---|---|

0

reset

<< Previous Page Next Page >>

#### Low Blood Sugar (Hypoglycemia)

Low blood sugar occurs when **blood sugar levels drop below the normal range**, causing symptoms such as dizziness, weakness, sweating, and confusion. While there are many possible causes, medication-induced low blood sugar is typically mild to moderate and can often be managed with simple dietary adjustments or fast-acting carbohydrates, such as sugar, for immediate relief.

* must provide value

| Not at all concerning 0 | Moderate Heart Attack 50 | Severe Disabling Stroke 100 |
|---|---|---|

0

reset

<< Previous Page Next Page >>

LOOBESITY ZURICH Universität Zürich USZ Universitäts Spital Zürich THE LOOP ZURICH

AAA English

**Hair Loss (Alopecia)**

Hair loss refers **to the partial or complete loss of hair.** While there are many potential causes, it is often temporary and caused by metabolic stress on the body, such as rapid weight loss, certain medications, trauma, surgery, chronic conditions, or childbirth. This occurs when hair follicles enter a resting phase, leading to increased shedding. In most cases, hair regrows once the metabolism stabilizes. However, in some cases, discontinuing the triggering medication may be considered, although this should be carefully evaluated if the medication is important, such as for weight reduction.

* must provide value

Not at all concerning 0 | Moderate Heart Attack 50 | Severe Disabling Stroke 100

0

reset

<< Previous Page | Next Page >>

**Gallstones (Cholelithiasis)**

Gallstones are **small, hard stones that form in the gallbladder or along the bile duct**. They can be caused by factors such as rapid weight loss, including that induced by weight management medications. While often asymptomatic, gallstones may sometimes cause mild to moderate intermittent pain, nausea, or digestive issues. Medications for dissolving the stones may be provided when necessary. In rare cases, severe symptoms can occur if the stones significantly block the bile ducts, requiring removal through surgical or non-surgical methods.

* must provide value

Not at all concerning 0 | Moderate Heart Attack 50 | Severe Disabling Stroke 100

0

reset

<< Previous Page | Next Page >>

**Fatigue**

Fatigue refers to a general **sense of tiredness and lack of energy** that can interfere with daily activities. Symptoms may include feeling weak, tired, and worn out, with severity often ranging from mild to moderate. Fatigue can have many causes, including certain medications. In most cases, it can be managed with lifestyle changes, such as maintaining good sleep hygiene, avoiding alcohol, adopting healthy eating habits, exercising regularly, or discontinuing medications that trigger it.

* must provide value

Not at all concerning 0 | Moderate Heart Attack 50 | Severe Disabling Stroke 100

0

reset

<< Previous Page | Next Page >>

**Constipation**

Constipation occurs when **stool is hard to pass**, often due to fewer bowel movements caused by certain medications or dietary factors. In most cases, it is mild and short-lived and can be managed at home by adding fiber-rich foods to your diet and staying hydrated. If needed, over-the-counter medications may help alleviate symptoms, or you might need to stop the medications that are causing it.

* must provide value

Not at all concerning 0 | Moderate Heart Attack 50 | Severe Disabling Stroke 100

0

reset

<< Previous Page | Next Page >>

**Upper Respiratory Tract Infection**

Upper respiratory tract infections affect the nose, throat, or sinuses and include conditions such as the **common cold, sore throat, and sinus infections**. Symptoms may include sore throat, fever, cough, and fatigue. These infections are often managed at home with rest, adequate hydration, and, if needed, over-the-counter medications to alleviate symptoms.

* must provide value

Not at all concerning 0 | Moderate Heart Attack 50 | Severe Disabling Stroke 100

0

reset

<< Previous Page | Next Page >>

LOOBESITY ZURICH | Universität Zürich | USZ Universitäts Spital Zürich | THE LOOP ZURICH

AAA

English

Page 4 of 22

##### Upper Abdominal Pain

Unlike abdominal pain, which can occur anywhere in the abdominal area, upper abdominal pain is a feeling of discomfort or **pain specifically in the upper part of your abdomen** (between your ribs and belly button). This pain often presents as mild and sometimes as moderate but can be alleviated by over-the-counter pain medications.

* must provide value

Not at all concerning 0 | Moderate Heart Attack 50 | Severe Disabling Stroke 100

0

reset

<< Previous Page    Next Page >>

##### Diarrhea

Diarrhea is characterized by **loose or watery stool and increased bowel movements**. It can be caused by certain medications or the quality and type of diet. While diarrhea can affect your daily activities, it can often be managed with lifestyle adjustments such as staying well-hydrated, modifying your diet, and avoiding foods or medications that trigger it. Over-the-counter medications may also help alleviate symptoms.

* must provide value

Not at all concerning 0 | Moderate Heart Attack 50 | Severe Disabling Stroke 100

0

reset

<< Previous Page    Next Page >>

##### Indigestion (Dyspepsia)

Indigestion is characterized **by pain or discomfort in the upper abdomen**, typically occurring after eating or as a side effect of certain medications, such as weight management drugs that slow gastric emptying. It is often accompanied by bloating or a feeling of fullness. Indigestion is commonly managed with lifestyle changes, such as modifying your diet, eating smaller meals, or stopping medications that trigger symptoms. In some cases, alternative therapies like acupuncture may be used, and over-the-counter medications can also help relieve symptoms.

* must provide value

Not at all concerning 0 | Moderate Heart Attack 50 | Severe Disabling Stroke 100

0

reset

<< Previous Page    Next Page >>

##### Vomiting

Unlike nausea, which is only a feeling, vomiting is the **forceful emptying of stomach contents** through the mouth and can have multiple causes. Like nausea, it often has many causes and is usually mild, but in some cases, it may significantly limit daily activities. Vomiting can be managed by sipping clear, cold fluids, consuming light and bland foods, and avoiding rich or greasy foods. Over-the-counter medications may also provide relief if needed.

* must provide value

Not at all concerning 0 | Moderate Heart Attack 50 | Severe Disabling Stroke 100

0

reset

<< Previous Page    Next Page >>

##### Nausea

Nausea is a feeling of **discomfort in the stomach that is often accompanied with the urge to vomit.** While often mild, it can disrupt your daily activities. Nausea can be alleviated by home remedies such as sipping fluids, consuming light, bland foods and avoiding rich or greasy foods. Also, some over-the-counter anti-nausea medications may provide relief if needed.

* must provide value

Not at all concerning 0 | Moderate Heart Attack 50 | Severe Disabling Stroke 100

0

reset

<< Previous Page    Next Page >>

##### Eructation (Belching/Burping)

Eructation, commonly known as belching or burping, refers to the **release of gas from the stomach or esophagus** through the mouth. It occurs when excess air is swallowed and becomes trapped in the esophagus or upper stomach. In some cases, it may be caused by certain medications. Eructation can often be minimized by avoiding carbonated drinks, chewing gum, and hard candies, eating slowly, or discontinuing medications that trigger it.

| Not at all concerning 0 | Moderate Heart Attack 50 | Severe Disabling Stroke 100 |
|---|---|---|

0

reset

* must provide value

<< Previous Page | Next Page >>

##### Flatulence

Flatulence is the **involuntary passing of gas** from the intestine through the rectum. It can often be managed with lifestyle modifications, such as adjusting your diet and exercise habits or addressing triggering factors, including discontinuing certain medications.

| Not at all concerning 0 | Moderate Heart Attack 50 | Severe Disabling Stroke 100 |
|---|---|---|

0

reset

* must provide value

<< Previous Page | Next Page >>

##### Dizziness

Dizziness encompasses a range of sensations, including **feeling faint, lightheaded, disoriented, unsteady, or weak.** It is typically mild to moderate in severity and can often be alleviated by resting and avoiding physical activities that may increase the risk of falls or injury.

| Not at all concerning 0 | Moderate Heart Attack 50 | Severe Disabling Stroke 100 |
|---|---|---|

0

reset

* must provide value

<< Previous Page | Next Page >>

##### Injection Site Reactions

Injection site reactions are **caused by the body's response to medications or substances administered through an injection**. These reactions include redness, swelling, and pain at the injection site. They typically improve on their own over time without the need for treatment. If needed, hot or cold compresses, over-the-counter pain relievers, or soothing topical oils can be used to alleviate pain and reduce the reactions.

| Not at all concerning 0 | Moderate Heart Attack 50 | Severe Disabling Stroke 100 |
|---|---|---|

0

reset

* must provide value

<< Previous Page | Next Page >>

**Part III:**

LOOBESITY ZURICH Universität Zürich USZ Universitäts Spital Zürich THE LOOP ZURICH English

##### Instructions

In the following 23 questions, you will be presented with a set of 5 health outcomes from the 21 described in Part 2.

Your task is to select:

- **The outcome that you find most concerning from the list.**
- **The outcome that you find least concerning from the list.**

Please focus on how you personally feel about each outcome in the context of treatment decisions. There are no right or wrong answers.

**Important Notes:**

- You will see the same outcomes appear multiple times in different combinations. This repetition helps us better understand your preferences.

Page 1 of 23

Part 3 of 3

**What is the least concerning and most concerning outcome to you?**

| Most Concerning | Least Concerning |
|---|---|
| ○ Abdominal Pain | ○ Abdominal Pain |
| ○ Gallbladder Inflammation | ○ Gallbladder Inflammation |
| ○ Dizziness | ○ Dizziness |
| ○ Indigestion | ○ Indigestion |
| ○ Belching/Burping | ○ Belching/Burping |
| reset | reset |

**What is the least concerning and most concerning outcome to you?**

| Most Concerning | Least Concerning |
|---|---|
| ○ Abdominal Pain | ○ Abdominal Pain |
| ○ Gallstones | ○ Gallstones |
| ○ Constipation | ○ Constipation |
| ○ Inflammation of the Pancreas | ○ Inflammation of the Pancreas |
| ○ Vomiting | ○ Vomiting |
| reset | reset |

<< Previous Page    Next Page >>

**What is the least concerning and most concerning outcome to you?**

| Most Concerning | Least Concerning |
|---|---|
| ○ Gallbladder Inflammation | ○ Gallbladder Inflammation |
| ○ Fatigue | ○ Fatigue |
| ○ Low Blood Sugar | ○ Low Blood Sugar |
| ○ Nausea | ○ Nausea |
| ○ Inflammation of the Pancreas | ○ Inflammation of the Pancreas |
| reset | reset |

<< Previous Page    Next Page >>

**What is the least concerning and most concerning outcome to you?**

| Most Concerning | Least Concerning |
|---|---|
| ○ Weight gain of 10% | ○ Weight gain of 10% |
| ○ Constipation | ○ Constipation |
| ○ Dizziness | ○ Dizziness |
| ○ Fatigue | ○ Fatigue |
| ○ Headache | ○ Headache |
| reset | reset |

<< Previous Page    Next Page >>

**What is the least concerning and most concerning outcome to you?**

| Most Concerning | Least Concerning |
|---|---|
| ◯ Gallstones | ◯ Gallstones |
| ◯ Dizziness | ◯ Dizziness |
| ◯ Low Blood Sugar | ◯ Low Blood Sugar |
| ◯ Injection Site Reactions | ◯ Injection Site Reactions |
| ◯ Upper Respiratory Tract Infections | ◯ Upper Respiratory Tract Infections |
| reset | reset |

<< Previous Page | Next Page >>

**What is the least concerning and most concerning outcome to you?**

| Most Concerning | Least Concerning |
|---|---|
| ◯ Diarrhea | ◯ Diarrhea |
| ◯ Dizziness | ◯ Dizziness |
| ◯ Flatulence | ◯ Flatulence |
| ◯ Inflammation of the Pancreas | ◯ Inflammation of the Pancreas |
| ◯ Upper Abdominal Pain | ◯ Upper Abdominal Pain |
| reset | reset |

<< Previous Page | Next Page >>

**What is the least concerning and most concerning outcome to you?**

| Most Concerning | Least Concerning |
|---|---|
| ◯ Weight Gain of 5% | ◯ Weight Gain of 5% |
| ◯ Indigestion | ◯ Indigestion |
| ◯ Headache | ◯ Headache |
| ◯ Inflammation of the Pancreas | ◯ Inflammation of the Pancreas |
| ◯ Upper Respiratory Tract Infection | ◯ Upper Respiratory Tract Infection |
| reset | reset |

<< Previous Page | Next Page >>

**What is the least concerning and most concerning outcome to you?**

| Most Concerning | Least Concerning |
|---|---|
| ◯ Weight Gain of 10% | ◯ Weight Gain of 10% |
| ◯ Indigestion | ◯ Indigestion |
| ◯ Low Blood Sugar | ◯ Low Blood Sugar |
| ◯ Upper Abdominal Pain | ◯ Upper Abdominal Pain |
| ◯ Vomiting | ◯ Vomiting |
| reset | reset |

<< Previous Page | Next Page >>

**What is the least concerning and most concerning outcome to you?**

| Most Concerning | Least Concerning |
|---|---|
| ◯ Belching/Burping | ◯ Belching/Burping |
| ◯ Fatigue | ◯ Fatigue |
| ◯ Flatulence | ◯ Flatulence |
| ◯ Vomiting | ◯ Vomiting |
| ◯ Upper Respiratory Tract Infection | ◯ Upper Respiratory Tract Infection |
| reset | reset |

<< Previous Page | Next Page >>

**What is the least concerning and most concerning outcome to you?**

| Most Concerning | Least Concerning |
|---|---|
| ○ Hair Loss | ○ Hair Loss |
| ○ Gallstones | ○ Gallstones |
| ○ Diarrhea | ○ Diarrhea |
| ○ Indigestion | ○ Indigestion |
| ○ Fatigue | ○ Fatigue |
| reset | reset |

<< Previous Page    Next Page >>

**What is the least concerning and most concerning outcome to you?**

| Most Concerning | Least Concerning |
|---|---|
| ○ Weight Gain of 5% | ○ Weight Gain of 5% |
| ○ Constipation | ○ Constipation |
| ○ Diarrhea | ○ Diarrhea |
| ○ Belching/Burping | ○ Belching/Burping |
| ○ Low Blood Sugar | ○ Low Blood Sugar |
| reset | reset |

<< Previous Page    Next Page >>

**What is the least concerning and most concerning outcome to you?**

| Most Concerning | Least Concerning |
|---|---|
| ○ Weight Gain of 5% | ○ Weight Gain of 5% |
| ○ Weight Gain of 10% | ○ Weight Gain of 10% |
| ○ Gallbladder Inflammation | ○ Gallbladder Inflammation |
| ○ Gallstones | ○ Gallstones |
| ○ Flatulence | ○ Flatulence |
| reset | reset |

<< Previous Page    Next Page >>

**What is the least concerning and most concerning outcome to you?**

| Most Concerning | Least Concerning |
|---|---|
| ○ Hair Loss | ○ Hair Loss |
| ○ Gallbladder Inflammation | ○ Gallbladder Inflammation |
| ○ Constipation | ○ Constipation |
| ○ Upper Abdominal Pain | ○ Upper Abdominal Pain |
| ○ Upper Respiratory Tract Infection | ○ Upper Respiratory Tract Infection |
| reset | reset |

<< Previous Page    Next Page >>

**What is the least concerning and most concerning outcome to you?**

| Most Concerning | Least Concerning |
|---|---|
| ○ Weight Gain of 10% | ○ Weight Gain of 10% |
| ○ Hair Loss | ○ Hair Loss |
| ○ Belching/Burping | ○ Belching/Burping |
| ○ Injection Site Reactions | ○ Injection Site Reactions |
| ○ Inflammation of the Pancreas | ○ Inflammation of the Pancreas |
| reset | reset |

<< Previous Page    Next Page >>

**What is the least concerning and most concerning outcome to you?**

| Most Concerning | Least Concerning |
|---|---|
| ○ Abdominal Pain | ○ Abdominal Pain |
| ○ Hair Loss | ○ Hair Loss |
| ○ Flatulence | ○ Flatulence |
| ○ Headache | ○ Headache |
| ○ Low Blood Sugar | ○ Low Blood Sugar |
| reset | reset |

< < Previous Page    Next Page >>

**What is the least concerning and most concerning outcome to you?**

| Most Concerning | Least Concerning |
|---|---|
| ○ Gallstones | ○ Gallstones |
| ○ Belching/Burping | ○ Belching/Burping |
| ○ Headache | ○ Headache |
| ○ Nausea | ○ Nausea |
| ○ Upper Abdominal Pain | ○ Upper Abdominal Pain |
| reset | reset |

< < Previous Page    Next Page >>

**What is the least concerning and most concerning outcome to you?**

| Most Concerning | Least Concerning |
|---|---|
| ○ Weight Gain of 10% | ○ Weight Gain of 10% |
| ○ Abdominal Pain | ○ Abdominal Pain |
| ○ Diarrhea | ○ Diarrhea |
| ○ Nausea | ○ Nausea |
| ○ Upper Respiratory Tract Infection | ○ Upper Respiratory Tract Infection |
| reset | reset |

< < Previous Page    Next Page >>

**What is the least concerning and most concerning outcome to you?**

| Most Concerning | Least Concerning |
|---|---|
| ○ Hair Loss | ○ Hair Loss |
| ○ Injection Site Reactions | ○ Injection Site Reactions |
| ○ Inflammation of the Pancreas | ○ Inflammation of the Pancreas |
| ○ Weight Gain of 10% | ○ Weight Gain of 10% |
| ○ Belching/Burping | ○ Belching/Burping |
| reset | reset |

< < Previous Page    Submit

**What is the least concerning and most concerning outcome to you?**

| Most Concerning | Least Concerning |
|---|---|
| ○ Low Blood Sugar | ○ Low Blood Sugar |
| ○ Upper Respiratory Tract Infection | ○ Upper Respiratory Tract Infection |
| ○ Dizziness | ○ Dizziness |
| ○ Injection Site Reactions | ○ Injection Site Reactions |
| ○ Gallstones | ○ Gallstones |
| reset | reset |

< < Previous Page    Next Page >>

**What is the least concerning and most concerning outcome to you?**

| Most Concerning | Least Concerning |
|---|---|
| ◯ Weight Gain of 5% | ◯ Weight Gain of 5% |
| ◯ Abdominal Pain | ◯ Abdominal Pain |
| ◯ Fatigue | ◯ Fatigue |
| ◯ Injection Site Reactions | ◯ Injection Site Reactions |
| ◯ Upper Abdominal Pain | ◯ Upper Abdominal Pain |
| reset | reset |

<< Previous Page    Next Page >>

**What is the least concerning and most concerning outcome to you?**

| Most Concerning | Least Concerning |
|---|---|
| ◯ Weight Gain of 5% | ◯ Weight Gain of 5% |
| ◯ Hair Loss | ◯ Hair Loss |
| ◯ Dizziness | ◯ Dizziness |
| ◯ Nausea | ◯ Nausea |
| ◯ Vomiting | ◯ Vomiting |
| reset | reset |

<< Previous Page    Next Page >>

**What is the least concerning and most concerning outcome to you?**

| Most Concerning | Least Concerning |
|---|---|
| ◯ Gallbladder Inflammation | ◯ Gallbladder Inflammation |
| ◯ Diarrhea | ◯ Diarrhea |
| ◯ Headache | ◯ Headache |
| ◯ Injection Site Reactions | ◯ Injection Site Reactions |
| ◯ Vomiting | ◯ Vomiting |
| reset | reset |

<< Previous Page    Next Page >>

**What is the least concerning and most concerning outcome to you?**

| Most Concerning | Least Concerning |
|---|---|
| ◯ Constipation | ◯ Constipation |
| ◯ Indigestion | ◯ Indigestion |
| ◯ Flatulence | ◯ Flatulence |
| ◯ Injection Site Reactions | ◯ Injection Site Reactions |
| ◯ Nausea | ◯ Nausea |
| reset | reset |

<< Previous Page    Next Page >>

#### Table S1: Subgroup-specific preference weights for each outcome

| Outcome | Sex: Female<br>Age: 18-29<br>BMI: 25-29<br>Exercise ≥150'/week: Yes | Female<br>18-29<br>25-29<br>No | Female<br>18-29<br>30-34<br>Yes | Female<br>18-29<br>30-34<br>No | Female<br>18-29<br>35+<br>Yes | Female<br>18-29<br>35+<br>No | Female<br>30-49<br>25-29<br>Yes | Female<br>30-49<br>25-29<br>No | Female<br>30-49<br>30-34<br>Yes |
|---|---|---|---|---|---|---|---|---|---|
| | **Number of Participants (n): 72** | **84** | **35** | **42** | **13** | **38** | **97** | **84** | **54** |
| Abdominal Pain | 0.18 (0.12 to 0.24) | 0.18 (0.12 to 0.24) | 0.18 (0.12 to 0.24) | 0.20 (0.14 to 0.27) | 0.21 (0.14 to 0.28) | 0.19 (0.13 to 0.26) | 0.21 (0.14 to 0.28) | 0.21 (0.14 to 0.29) | 0.20 (0.13 to 0.27) |
| Alopecia | 0.18 (0.12 to 0.28) | 0.18 (0.11 to 0.26) | 0.19 (0.11 to 0.29) | 0.19 (0.11 to 0.28) | 0.16 (0.09 to 0.26) | 0.16 (0.09 to 0.24) | 0.19 (0.11 to 0.27) | 0.18 (0.11 to 0.27) | 0.14 (0.08 to 0.22) |
| Constipation | 0.11 (0.07 to 0.16) | 0.11 (0.07 to 0.17) | 0.11 (0.07 to 0.16) | 0.11 (0.07 to 0.16) | 0.11 (0.07 to 0.17) | 0.11 (0.07 to 0.17) | 0.11 (0.07 to 0.16) | 0.11 (0.07 to 0.16) | 0.11 (0.07 to 0.17) |
| Diarrhea | 0.14 (0.09 to 0.19) | 0.14 (0.10 to 0.20) | 0.13 (0.09 to 0.19) | 0.14 (0.09 to 0.20) | 0.14 (0.09 to 0.19) | 0.14 (0.10 to 0.20) | 0.14 (0.09 to 0.20) | 0.14 (0.09 to 0.19) | 0.14 (0.10 to 0.20) |
| Dizziness | 0.18 (0.13 to 0.26) | 0.20 (0.14 to 0.27) | 0.18 (0.13 to 0.25) | 0.21 (0.15 to 0.29) | 0.20 (0.14 to 0.29) | 0.21 (0.16 to 0.30) | 0.20 (0.15 to 0.27) | 0.22 (0.16 to 0.30) | 0.21 (0.15 to 0.28) |
| Eructation | 0.04 (0.02 to 0.08) | 0.04 (0.02 to 0.08) | 0.04 (0.02 to 0.07) | 0.04 (0.02 to 0.08) | 0.04 (0.02 to 0.08) | 0.04 (0.02 to 0.08) | 0.04 (0.02 to 0.08) | 0.04 (0.02 to 0.08) | 0.04 (0.02 to 0.08) |
| Fatigue | 0.13 (0.08 to 0.19) | 0.13 (0.09 to 0.19) | 0.12 (0.08 to 0.18) | 0.13 (0.09 to 0.20) | 0.13 (0.09 to 0.20) | 0.13 (0.09 to 0.20) | 0.13 (0.09 to 0.19) | 0.12 (0.08 to 0.18) | 0.13 (0.08 to 0.19) |
| Flatulence | 0.05 (0.03 to 0.09) | 0.05 (0.03 to 0.09) | 0.05 (0.02 to 0.09) | 0.05 (0.03 to 0.09) | 0.05 (0.03 to 0.09) | 0.05 (0.03 to 0.09) | 0.05 (0.03 to 0.09) | 0.05 (0.03 to 0.09) | 0.05 (0.03 to 0.09) |
| Gallbladder Inflammation | 0.35 (0.26 to 0.46) | 0.38 (0.28 to 0.49) | 0.35 (0.26 to 0.45) | 0.36 (0.27 to 0.46) | 0.37 (0.28 to 0.48) | 0.35 (0.26 to 0.44) | 0.37 (0.27 to 0.48) | 0.37 (0.28 to 0.48) | 0.35 (0.26 to 0.45) |
| Gallstones | 0.31 (0.23 to 0.41) | 0.33 (0.24 to 0.42) | 0.32 (0.23 to 0.42) | 0.33 (0.24 to 0.43) | 0.34 (0.25 to 0.45) | 0.34 (0.25 to 0.44) | 0.34 (0.24 to 0.43) | 0.34 (0.25 to 0.44) | 0.31 (0.23 to 0.41) |
| Headache | 0.12 (0.08 to 0.19) | 0.12 (0.08 to 0.19) | 0.12 (0.08 to 0.18) | 0.12 (0.08 to 0.19) | 0.12 (0.08 to 0.19) | 0.13 (0.09 to 0.19) | 0.13 (0.08 to 0.19) | 0.12 (0.08 to 0.19) | 0.13 (0.09 to 0.19) |
| Hypoglycemia | 0.19 (0.14 to 0.27) | 0.19 (0.14 to 0.27) | 0.19 (0.13 to 0.26) | 0.19 (0.14 to 0.27) | 0.19 (0.14 to 0.27) | 0.20 (0.14 to 0.27) | 0.19 (0.13 to 0.27) | 0.19 (0.14 to 0.26) | 0.19 (0.14 to 0.27) |
| Indigestion | 0.13 (0.09 to 0.19) | 0.14 (0.09 to 0.20) | 0.14 (0.09 to 0.20) | 0.12 (0.08 to 0.18) | 0.12 (0.08 to 0.18) | 0.12 (0.08 to 0.18) | 0.11 (0.07 to 0.16) | 0.12 (0.08 to 0.18) | 0.12 (0.08 to 0.17) |
| Injection Site Reactions | 0.13 (0.08 to 0.20) | 0.12 (0.07 to 0.18) | 0.15 (0.08 to 0.22) | 0.11 (0.06 to 0.17) | 0.12 (0.07 to 0.20) | 0.12 (0.07 to 0.18) | 0.09 (0.05 to 0.14) | 0.11 (0.06 to 0.16) | 0.11 (0.06 to 0.17) |
| Nausea | 0.12 (0.08 to 0.18) | 0.12 (0.08 to 0.18) | 0.13 (0.08 to 0.18) | 0.11 (0.07 to 0.17) | 0.12 (0.08 to 0.17) | 0.12 (0.08 to 0.17) | 0.12 (0.08 to 0.18) | 0.12 (0.07 to 0.17) | 0.11 (0.07 to 0.17) |
| Pancreatitis | 0.45 (0.32 to 0.58) | 0.44 (0.32 to 0.57) | 0.41 (0.30 to 0.54) | 0.43 (0.31 to 0.56) | 0.45 (0.33 to 0.58) | 0.41 (0.29 to 0.53) | 0.44 (0.31 to 0.56) | 0.46 (0.33 to 0.60) | 0.43 (0.32 to 0.55) |
| Upper Abdominal Pain | 0.23 (0.17 to 0.31) | 0.24 (0.17 to 0.31) | 0.24 (0.17 to 0.31) | 0.24 (0.18 to 0.31) | 0.24 (0.18 to 0.32) | 0.24 (0.18 to 0.32) | 0.24 (0.17 to 0.31) | 0.24 (0.17 to 0.31) | 0.25 (0.18 to 0.32) |

| **Outcome** | **Sex: Female<br>Age: 18-29<br>BMI: 25-29<br>Exercise ≥150'/week: Yes** | **Female<br>18-29<br>25-29<br>No** | **Female<br>18-29<br>30-34<br>Yes** | **Female<br>18-29<br>30-34<br>No** | **Female<br>18-29<br>35+<br>Yes** | **Female<br>18-29<br>35+<br>No** | **Female<br>30-49<br>25-29<br>Yes** | **Female<br>30-49<br>25-29<br>No** | **Female<br>30-49<br>30-34<br>Yes** |
|---|---|---|---|---|---|---|---|---|---|
| Upper Respiratory Tract Infection | 0.24 (0.15 to 0.32) | 0.23 (0.14 to 0.31) | 0.23 (0.14 to 0.31) | 0.22 (0.14 to 0.31) | 0.22 (0.14 to 0.30) | 0.22 (0.14 to 0.30) | 0.21 (0.13 to 0.29) | 0.21 (0.13 to 0.29) | 0.23 (0.14 to 0.32) |
| Vomiting | 0.18 (0.13 to 0.25) | 0.19 (0.13 to 0.25) | 0.18 (0.13 to 0.25) | 0.19 (0.13 to 0.25) | 0.19 (0.13 to 0.25) | 0.19 (0.13 to 0.26) | 0.18 (0.13 to 0.25) | 0.18 (0.13 to 0.25) | 0.19 (0.13 to 0.25) |
| Weight Gain Of 10% | 0.42 (0.31 to 0.55) | 0.39 (0.29 to 0.50) | 0.42 (0.32 to 0.56) | 0.41 (0.31 to 0.54) | 0.41 (0.31 to 0.54) | 0.40 (0.30 to 0.53) | 0.42 (0.32 to 0.56) | 0.39 (0.29 to 0.51) | 0.42 (0.31 to 0.55) |
| Weight Gain Of 5% | 0.26 (0.19 to 0.35) | 0.23 (0.17 to 0.32) | 0.28 (0.20 to 0.37) | 0.24 (0.17 to 0.33) | 0.26 (0.18 to 0.36) | 0.24 (0.18 to 0.34) | 0.25 (0.18 to 0.34) | 0.23 (0.17 to 0.32) | 0.27 (0.20 to 0.37) |
| **Outcome** | **Sex: Female<br>Age: 30-49<br>BMI: 30-34<br>Exercise ≥150'/week No** | **Female<br>30-49<br>35+<br>Yes** | **Female<br>30-49<br>35+<br>No** | **Female<br>50+<br>25-29<br>Yes** | **Female<br>50+<br>25-29<br>No** | **Female<br>50+<br>30-34<br>Yes** | **Female<br>50+<br>30-34<br>No** | **Female<br>50+<br>35+<br>Yes** | **Female<br>50+<br>35+<br>No** |
| Number of Participants | n= 89 | 33 | 113 | 48 | 54 | 39 | 51 | 22 | 47 |
| Abdominal Pain | 0.22 (0.15 to 0.29) | 0.21 (0.15 to 0.29) | 0.19 (0.13 to 0.26) | 0.22 (0.15 to 0.29) | 0.23 (0.15 to 0.30) | 0.20 (0.14 to 0.27) | 0.23 (0.15 to 0.30) | 0.22 (0.15 to 0.29) | 0.22 (0.15 to 0.29) |
| Alopecia | 0.17 (0.10 to 0.25) | 0.17 (0.10 to 0.26) | 0.17 (0.11 to 0.24) | 0.14 (0.08 to 0.22) | 0.15 (0.09 to 0.24) | 0.15 (0.09 to 0.23) | 0.15 (0.09 to 0.23) | 0.14 (0.08 to 0.23) | 0.15 (0.09 to 0.23) |
| Constipation | 0.11 (0.07 to 0.17) | 0.11 (0.07 to 0.17) | 0.11 (0.07 to 0.16) | 0.12 (0.07 to 0.17) | 0.12 (0.08 to 0.17) | 0.11 (0.07 to 0.17) | 0.12 (0.07 to 0.17) | 0.11 (0.07 to 0.17) | 0.12 (0.08 to 0.17) |
| Diarrhea | 0.14 (0.09 to 0.19) | 0.14 (0.09 to 0.19) | 0.13 (0.09 to 0.19) | 0.14 (0.10 to 0.20) | 0.15 (0.10 to 0.21) | 0.14 (0.10 to 0.20) | 0.15 (0.10 to 0.20) | 0.14 (0.09 to 0.20) | 0.14 (0.10 to 0.20) |
| Dizziness | 0.21 (0.16 to 0.29) | 0.22 (0.16 to 0.30) | 0.20 (0.15 to 0.28) | 0.24 (0.17 to 0.32) | 0.23 (0.17 to 0.31) | 0.24 (0.18 to 0.33) | 0.24 (0.18 to 0.33) | 0.24 (0.17 to 0.33) | 0.25 (0.18 to 0.34) |
| Eructation | 0.04 (0.02 to 0.08) | 0.04 (0.02 to 0.08) | 0.04 (0.02 to 0.08) | 0.04 (0.02 to 0.08) | 0.05 (0.02 to 0.08) | 0.04 (0.02 to 0.08) | 0.05 (0.02 to 0.08) | 0.05 (0.02 to 0.08) | 0.05 (0.02 to 0.08) |
| Fatigue | 0.13 (0.09 to 0.20) | 0.13 (0.09 to 0.19) | 0.12 (0.08 to 0.19) | 0.14 (0.09 to 0.20) | 0.14 (0.09 to 0.21) | 0.13 (0.09 to 0.20) | 0.14 (0.10 to 0.21) | 0.14 (0.09 to 0.21) | 0.14 (0.10 to 0.21) |
| Flatulence | 0.05 (0.03 to 0.09) | 0.05 (0.03 to 0.09) | 0.05 (0.02 to 0.09) | 0.05 (0.03 to 0.09) | 0.06 (0.03 to 0.09) | 0.05 (0.03 to 0.09) | 0.05 (0.03 to 0.09) | 0.05 (0.03 to 0.09) | 0.05 (0.03 to 0.09) |
| Gallbladder Inflammation | 0.35 (0.26 to 0.45) | 0.34 (0.26 to 0.44) | 0.37 (0.28 to 0.49) | 0.36 (0.26 to 0.47) | 0.35 (0.26 to 0.45) | 0.34 (0.25 to 0.44) | 0.34 (0.25 to 0.44) | 0.36 (0.27 to 0.47) | 0.35 (0.26 to 0.45) |
| Gallstones | 0.31 (0.23 to 0.41) | 0.33 (0.24 to 0.42) | 0.34 (0.25 to 0.45) | 0.33 (0.24 to 0.43) | 0.32 (0.23 to 0.41) | 0.30 (0.22 to 0.40) | 0.31 (0.23 to 0.41) | 0.31 (0.22 to 0.40) | 0.33 (0.24 to 0.43) |
| Headache | 0.13 (0.09 to 0.20) | 0.13 (0.08 to 0.19) | 0.13 (0.09 to 0.20) | 0.14 (0.09 to 0.21) | 0.14 (0.09 to 0.21) | 0.13 (0.09 to 0.20) | 0.14 (0.09 to 0.21) | 0.13 (0.09 to 0.20) | 0.13 (0.09 to 0.20) |

| Outcome | Sex: Female<br>Age: 18-29<br>BMI: 25-29<br>Exercise ≥150'/week: Yes | Female<br>18-29<br>25-29<br>No | Female<br>18-29<br>30-34<br>Yes | Female<br>18-29<br>30-34<br>No | Female<br>18-29<br>35+<br>Yes | Female<br>18-29<br>35+<br>No | Female<br>30-49<br>25-29<br>Yes | Female<br>30-49<br>25-29<br>No | Female<br>30-49<br>30-34<br>Yes |
|---|---|---|---|---|---|---|---|---|---|
| Hypoglycemia | 0.19 (0.14 to 0.27) | 0.19 (0.14 to 0.27) | 0.19 (0.14 to 0.27) | 0.20 (0.14 to 0.27) | 0.20 (0.14 to 0.28) | 0.19 (0.14 to 0.27) | 0.20 (0.14 to 0.27) | 0.20 (0.14 to 0.27) | 0.20 (0.14 to 0.28) |
| Indigestion | 0.11 (0.08 to 0.16) | 0.12 (0.08 to 0.17) | 0.11 (0.07 to 0.16) | 0.12 (0.08 to 0.17) | 0.13 (0.08 to 0.18) | 0.12 (0.08 to 0.17) | 0.12 (0.08 to 0.17) | 0.12 (0.08 to 0.18) | 0.12 (0.08 to 0.18) |
| Injection Site Reactions | 0.12 (0.07 to 0.18) | 0.10 (0.05 to 0.16) | 0.09 (0.05 to 0.14) | 0.06 (0.03 to 0.11) | 0.08 (0.04 to 0.13) | 0.09 (0.05 to 0.14) | 0.07 (0.04 to 0.12) | 0.09 (0.04 to 0.15) | 0.08 (0.04 to 0.13) |
| Nausea | 0.12 (0.08 to 0.17) | 0.12 (0.08 to 0.17) | 0.11 (0.07 to 0.16) | 0.13 (0.08 to 0.19) | 0.12 (0.08 to 0.18) | 0.13 (0.08 to 0.18) | 0.12 (0.08 to 0.18) | 0.13 (0.08 to 0.19) | 0.12 (0.08 to 0.18) |
| Pancreatitis | 0.42 (0.30 to 0.55) | 0.44 (0.32 to 0.57) | 0.47 (0.35 to 0.60) | 0.47 (0.34 to 0.61) | 0.45 (0.33 to 0.58) | 0.44 (0.32 to 0.58) | 0.42 (0.30 to 0.54) | 0.43 (0.30 to 0.56) | 0.42 (0.30 to 0.54) |
| Upper Abdominal Pain | 0.24 (0.17 to 0.31) | 0.24 (0.18 to 0.32) | 0.24 (0.17 to 0.31) | 0.25 (0.18 to 0.33) | 0.26 (0.19 to 0.34) | 0.24 (0.17 to 0.32) | 0.25 (0.18 to 0.33) | 0.25 (0.18 to 0.32) | 0.25 (0.18 to 0.33) |
| Upper Respiratory Tract Infection | 0.22 (0.14 to 0.31) | 0.21 (0.13 to 0.30) | 0.24 (0.15 to 0.33) | 0.22 (0.14 to 0.31) | 0.23 (0.14 to 0.32) | 0.22 (0.14 to 0.31) | 0.22 (0.14 to 0.31) | 0.22 (0.14 to 0.31) | 0.22 (0.14 to 0.30) |
| Vomiting | 0.19 (0.13 to 0.25) | 0.19 (0.13 to 0.25) | 0.18 (0.13 to 0.25) | 0.19 (0.13 to 0.26) | 0.19 (0.13 to 0.26) | 0.19 (0.13 to 0.26) | 0.19 (0.13 to 0.26) | 0.19 (0.13 to 0.25) | 0.19 (0.13 to 0.26) |
| Weight Gain Of 10% | 0.39 (0.29 to 0.51) | 0.41 (0.31 to 0.54) | 0.43 (0.33 to 0.56) | 0.39 (0.29 to 0.53) | 0.40 (0.30 to 0.52) | 0.41 (0.31 to 0.54) | 0.41 (0.31 to 0.54) | 0.41 (0.31 to 0.55) | 0.43 (0.32 to 0.56) |
| Weight Gain Of 5% | 0.24 (0.17 to 0.32) | 0.25 (0.18 to 0.35) | 0.28 (0.21 to 0.39) | 0.20 (0.14 to 0.28) | 0.20 (0.15 to 0.29) | 0.25 (0.18 to 0.35) | 0.24 (0.17 to 0.33) | 0.24 (0.17 to 0.34) | 0.26 (0.19 to 0.35) |
| **Outcome** | **Sex: Male<br>Age: 18-29<br>BMI: 25-29<br>Exercise ≥150'/week<br>Yes** | **Male<br>18-29<br>25-29<br>No** | **Male<br>18-29<br>30-34<br>Yes** | **Male<br>18-29<br>30-34<br>No** | **Male<br>18-29<br>35+<br>Yes** | **Male<br>18-29<br>35+<br>No** | **Male<br>30-49<br>25-29<br>Yes** | **Male<br>30-49<br>25-29<br>No** | **Male<br>30-49<br>30-34<br>Yes** |
| Number of Participants | n= 117 | 73 | 33 | 30 | 15 | 16 | 168 | 139 | 72 |
| Abdominal Pain | 0.18 (0.12 to 0.24) | 0.18 (0.12 to 0.25) | 0.21 (0.14 to 0.28) | 0.19 (0.13 to 0.26) | 0.19 (0.12 to 0.26) | 0.18 (0.12 to 0.26) | 0.19 (0.13 to 0.25) | 0.20 (0.13 to 0.26) | 0.21 (0.14 to 0.27) |
| Alopecia | 0.14 (0.09 to 0.22) | 0.16 (0.10 to 0.23) | 0.15 (0.09 to 0.23) | 0.16 (0.09 to 0.25) | 0.11 (0.05 to 0.18) | 0.15 (0.08 to 0.24) | 0.13 (0.08 to 0.19) | 0.12 (0.07 to 0.19) | 0.15 (0.09 to 0.22) |
| Constipation | 0.11 (0.07 to 0.17) | 0.11 (0.07 to 0.17) | 0.11 (0.07 to 0.17) | 0.11 (0.07 to 0.17) | 0.11 (0.07 to 0.17) | 0.11 (0.07 to 0.17) | 0.11 (0.08 to 0.17) | 0.11 (0.07 to 0.17) | 0.11 (0.07 to 0.17) |
| Diarrhea | 0.14 (0.09 to 0.19) | 0.14 (0.10 to 0.20) | 0.14 (0.09 to 0.20) | 0.14 (0.09 to 0.20) | 0.14 (0.10 to 0.20) | 0.14 (0.09 to 0.20) | 0.14 (0.10 to 0.20) | 0.14 (0.10 to 0.20) | 0.14 (0.09 to 0.19) |

| Outcome | Sex: Female<br>Age: 18-29<br>BMI: 25-29<br>Exercise ≥150’/week: Yes | Female<br>18-29<br>25-29<br>No | Female<br>18-29<br>30-34<br>Yes | Female<br>18-29<br>30-34<br>No | Female<br>18-29<br>35+<br>Yes | Female<br>18-29<br>35+<br>No | Female<br>30-49<br>25-29<br>Yes | Female<br>30-49<br>25-29<br>No | Female<br>30-49<br>30-34<br>Yes |
|---|---|---|---|---|---|---|---|---|---|
| Dizziness | 0.18 (0.13 to 0.26) | 0.19 (0.14 to 0.26) | 0.18 (0.13 to 0.26) | 0.20 (0.14 to 0.28) | 0.18 (0.13 to 0.27) | 0.20 (0.14 to 0.29) | 0.21 (0.16 to 0.29) | 0.21 (0.15 to 0.29) | 0.22 (0.16 to 0.30) |
| Eructation | 0.04 (0.02 to 0.08) | 0.04 (0.02 to 0.08) | 0.04 (0.02 to 0.08) | 0.04 (0.02 to 0.08) | 0.04 (0.02 to 0.08) | 0.04 (0.02 to 0.08) | 0.05 (0.02 to 0.08) | 0.04 (0.02 to 0.08) | 0.04 (0.02 to 0.08) |
| Fatigue | 0.13 (0.08 to 0.19) | 0.13 (0.09 to 0.19) | 0.13 (0.09 to 0.20) | 0.13 (0.09 to 0.20) | 0.13 (0.09 to 0.20) | 0.13 (0.09 to 0.20) | 0.13 (0.09 to 0.19) | 0.13 (0.09 to 0.20) | 0.13 (0.08 to 0.19) |
| Flatulence | 0.05 (0.03 to 0.09) | 0.05 (0.03 to 0.09) | 0.05 (0.03 to 0.09) | 0.05 (0.03 to 0.09) | 0.05 (0.03 to 0.09) | 0.05 (0.02 to 0.09) | 0.05 (0.03 to 0.09) | 0.05 (0.03 to 0.09) | 0.05 (0.03 to 0.09) |
| Gallbladder Inflammation | 0.36 (0.27 to 0.46) | 0.35 (0.26 to 0.45) | 0.34 (0.25 to 0.44) | 0.35 (0.26 to 0.46) | 0.36 (0.27 to 0.48) | 0.36 (0.27 to 0.46) | 0.34 (0.25 to 0.44) | 0.34 (0.25 to 0.44) | 0.34 (0.25 to 0.45) |
| Gallstones | 0.31 (0.23 to 0.41) | 0.32 (0.23 to 0.41) | 0.34 (0.25 to 0.44) | 0.33 (0.24 to 0.43) | 0.33 (0.23 to 0.44) | 0.33 (0.24 to 0.43) | 0.32 (0.23 to 0.41) | 0.33 (0.24 to 0.43) | 0.30 (0.22 to 0.40) |
| Headache | 0.13 (0.09 to 0.20) | 0.13 (0.09 to 0.20) | 0.12 (0.08 to 0.19) | 0.12 (0.08 to 0.20) | 0.13 (0.09 to 0.20) | 0.12 (0.08 to 0.20) | 0.13 (0.09 to 0.20) | 0.13 (0.09 to 0.20) | 0.13 (0.08 to 0.19) |
| Hypoglycemia | 0.19 (0.14 to 0.27) | 0.19 (0.14 to 0.27) | 0.19 (0.14 to 0.27) | 0.19 (0.14 to 0.27) | 0.20 (0.14 to 0.27) | 0.19 (0.14 to 0.27) | 0.19 (0.14 to 0.27) | 0.19 (0.14 to 0.27) | 0.19 (0.13 to 0.26) |
| Indigestion | 0.14 (0.09 to 0.20) | 0.14 (0.09 to 0.19) | 0.14 (0.09 to 0.20) | 0.13 (0.09 to 0.19) | 0.14 (0.09 to 0.20) | 0.12 (0.08 to 0.18) | 0.12 (0.08 to 0.18) | 0.13 (0.08 to 0.19) | 0.12 (0.08 to 0.18) |
| Injection Site Reactions | 0.15 (0.09 to 0.22) | 0.15 (0.09 to 0.22) | 0.16 (0.09 to 0.24) | 0.12 (0.06 to 0.18) | 0.14 (0.08 to 0.21) | 0.13 (0.07 to 0.21) | 0.12 (0.07 to 0.18) | 0.11 (0.06 to 0.16) | 0.13 (0.07 to 0.18) |
| Nausea | 0.13 (0.08 to 0.18) | 0.13 (0.09 to 0.19) | 0.12 (0.08 to 0.18) | 0.13 (0.08 to 0.19) | 0.13 (0.08 to 0.19) | 0.13 (0.08 to 0.19) | 0.13 (0.09 to 0.19) | 0.13 (0.09 to 0.19) | 0.13 (0.08 to 0.18) |
| Pancreatitis | 0.40 (0.29 to 0.53) | 0.42 (0.31 to 0.55) | 0.37 (0.26 to 0.49) | 0.40 (0.28 to 0.53) | 0.43 (0.31 to 0.56) | 0.43 (0.32 to 0.57) | 0.40 (0.29 to 0.52) | 0.42 (0.30 to 0.54) | 0.40 (0.29 to 0.52) |
| Upper Abdominal Pain | 0.24 (0.17 to 0.31) | 0.24 (0.18 to 0.31) | 0.24 (0.17 to 0.32) | 0.24 (0.17 to 0.32) | 0.25 (0.18 to 0.32) | 0.25 (0.18 to 0.32) | 0.24 (0.17 to 0.31) | 0.25 (0.18 to 0.32) | 0.24 (0.17 to 0.31) |
| Upper Respiratory Tract Infection | 0.24 (0.15 to 0.34) | 0.22 (0.14 to 0.30) | 0.22 (0.14 to 0.31) | 0.23 (0.14 to 0.31) | 0.23 (0.15 to 0.33) | 0.22 (0.14 to 0.32) | 0.22 (0.13 to 0.30) | 0.22 (0.14 to 0.31) | 0.22 (0.14 to 0.31) |
| Vomiting | 0.18 (0.13 to 0.25) | 0.19 (0.13 to 0.25) | 0.18 (0.13 to 0.25) | 0.19 (0.13 to 0.26) | 0.19 (0.13 to 0.26) | 0.19 (0.13 to 0.26) | 0.19 (0.13 to 0.26) | 0.19 (0.13 to 0.26) | 0.18 (0.13 to 0.25) |
| Weight Gain Of 10% | 0.39 (0.29 to 0.51) | 0.40 (0.30 to 0.52) | 0.42 (0.32 to 0.56) | 0.41 (0.30 to 0.53) | 0.41 (0.31 to 0.54) | 0.42 (0.32 to 0.56) | 0.43 (0.33 to 0.55) | 0.41 (0.31 to 0.54) | 0.41 (0.31 to 0.53) |
| Weight Gain Of 5% | 0.21 (0.15 to 0.29) | 0.21 (0.15 to 0.29) | 0.25 (0.18 to 0.35) | 0.23 (0.17 to 0.32) | 0.25 (0.17 to 0.35) | 0.27 (0.19 to 0.38) | 0.23 (0.17 to 0.32) | 0.24 (0.18 to 0.32) | 0.25 (0.18 to 0.34) |
| **Outcome** | **Sex: Male<br>Age: 30-49<br>BMI: 30-34** | **Male<br>30-49** | **Male<br>30-49** | **Male<br>50+** | **Male<br>50+** | **Male<br>50+** | **Male<br>50+** | **Male<br>50+** | **Male<br>50+** |

| Outcome | Sex: Female<br>Age: 18-29<br>BMI: 25-29<br>Exercise ≥150’/week: Yes | Female<br>18-29<br>25-29<br>No | Female<br>18-29<br>30-34<br>Yes | Female<br>18-29<br>30-34<br>No | Female<br>18-29<br>35+<br>Yes | Female<br>18-29<br>35+<br>No | Female<br>30-49<br>25-29<br>Yes | Female<br>30-49<br>25-29<br>No | Female<br>30-49<br>30-34<br>Yes |
|---|---|---|---|---|---|---|---|---|---|
| | **Exercise ≥150’/week No** | **35+ Yes** | **35+ No** | **25-29 Yes** | **25-29 No** | **30-34 Yes** | **30-34 No** | **35+ Yes** | **35+ No** |
| Number of Participants | n= 83 | 27 | 69 | 53 | 60 | 37 | 58 | 16 | 31 |
| Abdominal Pain | 0.20 (0.13 to 0.27) | 0.23 (0.15 to 0.30) | 0.22 (0.15 to 0.30) | 0.23 (0.15 to 0.31) | 0.23 (0.16 to 0.31) | 0.22 (0.14 to 0.29) | 0.23 (0.16 to 0.31) | 0.22 (0.14 to 0.30) | 0.20 (0.14 to 0.28) |
| Alopecia | 0.14 (0.08 to 0.21) | 0.09 (0.04 to 0.15) | 0.11 (0.07 to 0.17) | 0.10 (0.06 to 0.16) | 0.09 (0.05 to 0.15) | 0.08 (0.04 to 0.14) | 0.09 (0.05 to 0.14) | 0.10 (0.05 to 0.17) | 0.09 (0.05 to 0.15) |
| Constipation | 0.11 (0.07 to 0.17) | 0.11 (0.07 to 0.17) | 0.11 (0.07 to 0.16) | 0.11 (0.07 to 0.17) | 0.12 (0.08 to 0.18) | 0.12 (0.08 to 0.18) | 0.12 (0.08 to 0.17) | 0.12 (0.08 to 0.18) | 0.11 (0.07 to 0.17) |
| Diarrhea | 0.14 (0.09 to 0.19) | 0.14 (0.10 to 0.20) | 0.14 (0.09 to 0.19) | 0.15 (0.10 to 0.21) | 0.15 (0.10 to 0.21) | 0.15 (0.10 to 0.21) | 0.14 (0.10 to 0.20) | 0.15 (0.10 to 0.22) | 0.14 (0.10 to 0.20) |
| Dizziness | 0.21 (0.15 to 0.28) | 0.22 (0.16 to 0.31) | 0.21 (0.15 to 0.29) | 0.24 (0.18 to 0.33) | 0.23 (0.17 to 0.32) | 0.23 (0.17 to 0.32) | 0.26 (0.19 to 0.36) | 0.23 (0.17 to 0.33) | 0.24 (0.17 to 0.33) |
| Eructation | 0.04 (0.02 to 0.08) | 0.04 (0.02 to 0.08) | 0.04 (0.02 to 0.08) | 0.04 (0.02 to 0.08) | 0.05 (0.02 to 0.08) | 0.05 (0.02 to 0.08) | 0.04 (0.02 to 0.08) | 0.05 (0.02 to 0.09) | 0.04 (0.02 to 0.08) |
| Fatigue | 0.13 (0.09 to 0.20) | 0.13 (0.09 to 0.20) | 0.13 (0.09 to 0.19) | 0.14 (0.09 to 0.20) | 0.14 (0.10 to 0.22) | 0.14 (0.10 to 0.21) | 0.14 (0.09 to 0.21) | 0.15 (0.10 to 0.22) | 0.13 (0.09 to 0.20) |
| Flatulence | 0.05 (0.03 to 0.09) | 0.05 (0.03 to 0.09) | 0.05 (0.03 to 0.09) | 0.05 (0.03 to 0.09) | 0.05 (0.03 to 0.09) | 0.06 (0.03 to 0.09) | 0.05 (0.03 to 0.09) | 0.06 (0.03 to 0.10) | 0.05 (0.03 to 0.09) |
| Gallbladder Inflammation | 0.36 (0.26 to 0.46) | 0.36 (0.26 to 0.47) | 0.34 (0.25 to 0.44) | 0.34 (0.26 to 0.44) | 0.34 (0.25 to 0.43) | 0.36 (0.27 to 0.46) | 0.35 (0.26 to 0.45) | 0.35 (0.26 to 0.47) | 0.33 (0.24 to 0.43) |
| Gallstones | 0.33 (0.24 to 0.42) | 0.32 (0.23 to 0.42) | 0.32 (0.23 to 0.42) | 0.32 (0.23 to 0.42) | 0.31 (0.23 to 0.41) | 0.35 (0.25 to 0.45) | 0.33 (0.24 to 0.43) | 0.32 (0.23 to 0.42) | 0.34 (0.25 to 0.44) |
| Headache | 0.12 (0.08 to 0.19) | 0.12 (0.08 to 0.20) | 0.12 (0.08 to 0.19) | 0.14 (0.09 to 0.21) | 0.14 (0.09 to 0.20) | 0.13 (0.09 to 0.20) | 0.13 (0.09 to 0.20) | 0.14 (0.10 to 0.22) | 0.13 (0.09 to 0.21) |
| Hypoglycemia | 0.19 (0.14 to 0.26) | 0.19 (0.14 to 0.27) | 0.19 (0.14 to 0.27) | 0.20 (0.14 to 0.27) | 0.20 (0.14 to 0.28) | 0.20 (0.14 to 0.29) | 0.20 (0.14 to 0.28) | 0.21 (0.15 to 0.29) | 0.20 (0.14 to 0.28) |
| Indigestion | 0.12 (0.08 to 0.17) | 0.12 (0.08 to 0.18) | 0.12 (0.08 to 0.17) | 0.12 (0.08 to 0.18) | 0.12 (0.08 to 0.18) | 0.12 (0.08 to 0.18) | 0.12 (0.08 to 0.18) | 0.12 (0.08 to 0.18) | 0.12 (0.07 to 0.17) |
| Injection Site Reactions | 0.12 (0.07 to 0.18) | 0.10 (0.05 to 0.16) | 0.12 (0.07 to 0.18) | 0.09 (0.05 to 0.15) | 0.09 (0.05 to 0.14) | 0.09 (0.05 to 0.15) | 0.09 (0.05 to 0.14) | 0.09 (0.05 to 0.15) | 0.09 (0.05 to 0.14) |
| Nausea | 0.12 (0.08 to 0.18) | 0.13 (0.08 to 0.18) | 0.12 (0.08 to 0.18) | 0.14 (0.09 to 0.20) | 0.14 (0.09 to 0.21) | 0.13 (0.09 to 0.19) | 0.13 (0.08 to 0.19) | 0.14 (0.09 to 0.21) | 0.13 (0.08 to 0.18) |
| Pancreatitis | 0.43 (0.31 to 0.55) | 0.44 (0.32 to 0.59) | 0.40 (0.29 to 0.52) | 0.43 (0.31 to 0.56) | 0.41 (0.29 to 0.54) | 0.45 (0.33 to 0.58) | 0.42 (0.30 to 0.55) | 0.42 (0.30 to 0.55) | 0.43 (0.31 to 0.56) |
| Upper Abdominal Pain | 0.24 (0.17 to 0.31) | 0.25 (0.18 to 0.32) | 0.24 (0.17 to 0.32) | 0.25 (0.18 to 0.33) | 0.26 (0.19 to 0.34) | 0.26 (0.19 to 0.33) | 0.26 (0.18 to 0.33) | 0.26 (0.18 to 0.34) | 0.25 (0.18 to 0.33) |

| **Outcome** | **Sex: Female**<br>**Age: 18-29**<br>**BMI: 25-29**<br>**Exercise ≥150'/week: Yes** | **Female**<br>**18-29**<br>**25-29**<br>**No** | **Female**<br>**18-29**<br>**30-34**<br>**Yes** | **Female**<br>**18-29**<br>**30-34**<br>**No** | **Female**<br>**18-29**<br>**35+**<br>**Yes** | **Female**<br>**18-29**<br>**35+**<br>**No** | **Female**<br>**30-49**<br>**25-29**<br>**Yes** | **Female**<br>**30-49**<br>**25-29**<br>**No** | **Female**<br>**30-49**<br>**30-34**<br>**Yes** |
|---|---|---|---|---|---|---|---|---|---|
| Upper Respiratory Tract Infection | 0.23 (0.14 to 0.31) | 0.23 (0.14 to 0.32) | 0.23 (0.14 to 0.32) | 0.23 (0.14 to 0.31) | 0.23 (0.14 to 0.31) | 0.23 (0.15 to 0.32) | 0.23 (0.14 to 0.31) | 0.24 (0.15 to 0.34) | 0.24 (0.15 to 0.33) |
| Vomiting | 0.19 (0.13 to 0.25) | 0.19 (0.13 to 0.26) | 0.19 (0.13 to 0.25) | 0.19 (0.13 to 0.26) | 0.20 (0.14 to 0.26) | 0.20 (0.14 to 0.27) | 0.19 (0.13 to 0.26) | 0.20 (0.14 to 0.27) | 0.19 (0.13 to 0.26) |
| Weight Gain Of 10% | 0.41 (0.30 to 0.53) | 0.43 (0.32 to 0.57) | 0.43 (0.32 to 0.55) | 0.40 (0.30 to 0.53) | 0.40 (0.29 to 0.53) | 0.41 (0.31 to 0.54) | 0.41 (0.30 to 0.54) | 0.43 (0.31 to 0.56) | 0.43 (0.32 to 0.55) |
| Weight Gain Of 5% | 0.25 (0.18 to 0.34) | 0.28 (0.20 to 0.39) | 0.28 (0.21 to 0.38) | 0.20 (0.15 to 0.29) | 0.22 (0.16 to 0.30) | 0.24 (0.17 to 0.33) | 0.24 (0.17 to 0.33) | 0.25 (0.18 to 0.35) | 0.26 (0.19 to 0.36) |

#### Table S2: $R^2$ for the subgroup model for estimating preference weights

| Subgroup (Sex\|Age\|BMI\|Exercise) | $R^2$ |
|---|---|
| Female \| 18-29 \| 25-29 \| Yes | 0.59 |
| Female \| 18-29 \| 25-29 \| No | 0.56 |
| Female \| 18-29 \| 30-34 \| Yes | 0.40 |
| Female \| 18-29 \| 30-34 \| No | 0.54 |
| Female \| 18-29 \| 35+ \| Yes | 0.55 |
| Female \| 18-29 \| 35+ \| No | 0.51 |
| Female \| 30-49 \| 25-29 \| Yes | 0.75 |
| Female \| 30-49 \| 25-29 \| No | 0.65 |
| Female \| 30-49 \| 30-34 \| Yes | 0.75 |
| Female \| 30-49 \| 30-34 \| No | 0.66 |
| Female \| 30-49 \| 35+ \| Yes | 0.74 |
| Female \| 30-49 \| 35+ \| No | 0.71 |
| Female \| 50+ \| 25-29 \| Yes | 0.65 |
| Female \| 50+ \| 25-29 \| No | 0.77 |
| Female \| 50+ \| 30-34 \| Yes | 0.71 |
| Female \| 50+ \| 30-34 \| No | 0.66 |
| Female \| 50+ \| 35+ \| Yes | 0.65 |
| Female \| 50+ \| 35+ \| No | 0.74 |
| Male \| 18-29 \| 25-29 \| Yes | 0.46 |
| Male \| 18-29 \| 25-29 \| No | 0.58 |
| Male \| 18-29 \| 30-34 \| Yes | 0.51 |
| Male \| 18-29 \| 30-34 \| No | 0.50 |
| Male \| 18-29 \| 35+ \| Yes | 0.32 |
| Male \| 18-29 \| 35+ \| No | 0.36 |
| Male \| 30-49 \| 25-29 \| Yes | 0.72 |
| Male \| 30-49 \| 25-29 \| No | 0.70 |
| Male \| 30-49 \| 30-34 \| Yes | 0.65 |
| Male \| 30-49 \| 30-34 \| No | 0.70 |
| Male \| 30-49 \| 35+ \| Yes | 0.47 |
| Male \| 30-49 \| 35+ \| No | 0.60 |
| Male \| 50+ \| 25-29 \| Yes | 0.61 |
| Male \| 50+ \| 25-29 \| No | 0.48 |
| Male \| 50+ \| 30-34 \| Yes | 0.78 |
| Male \| 50+ \| 30-34 \| No | 0.70 |
| Male \| 50+ \| 35+ \| Yes | 0.64 |
| Male \| 50+ \| 35+ \| No | 0.59 |

**Figure S1 Correlation plot between preference weights and visual analogue scale**

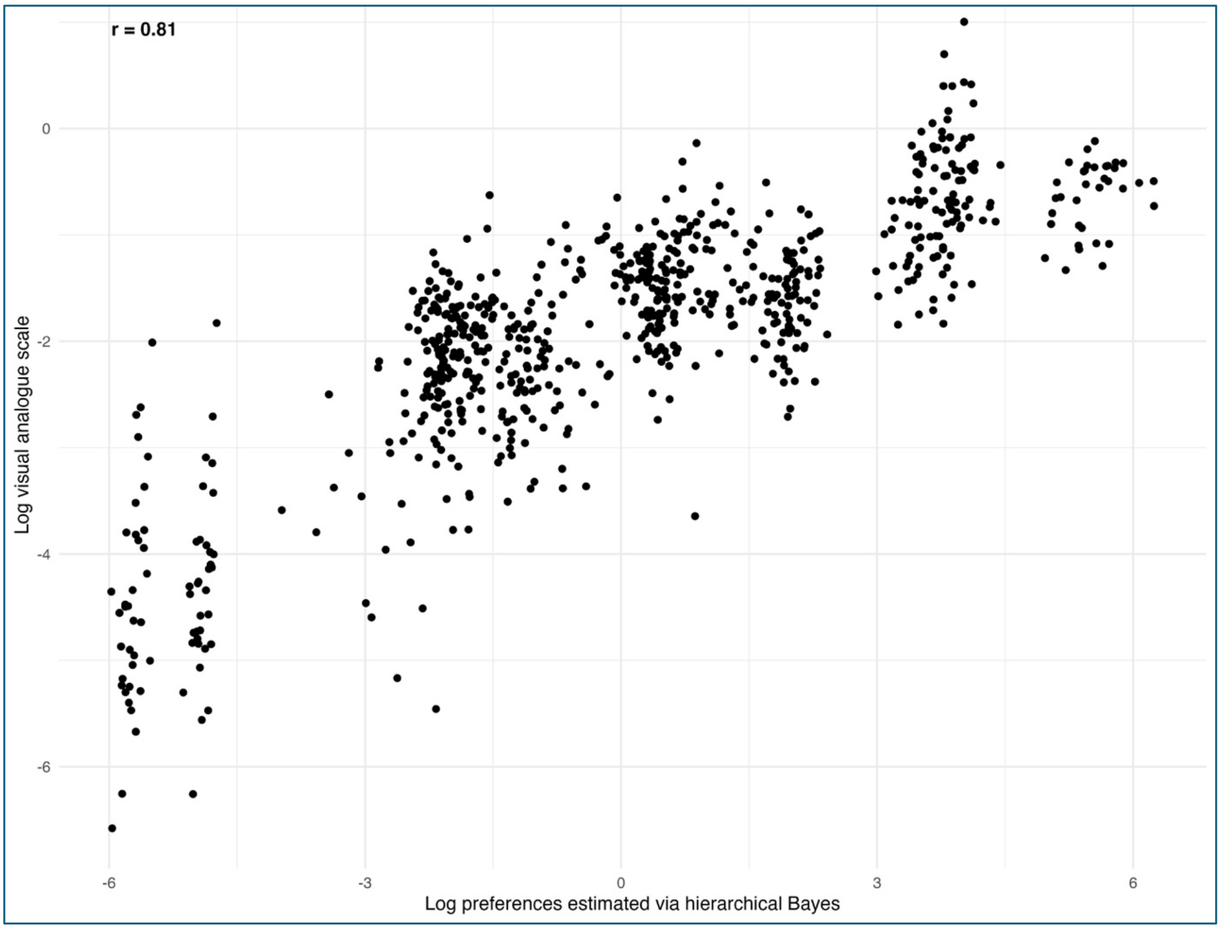

Each point represents the mean log-transformed values for a given outcome within a subgroup. The two measures were strongly correlated (r = 0.81, Pearson correlation coefficient).

#### Figure S2: Distribution of individual preference weights by outcome

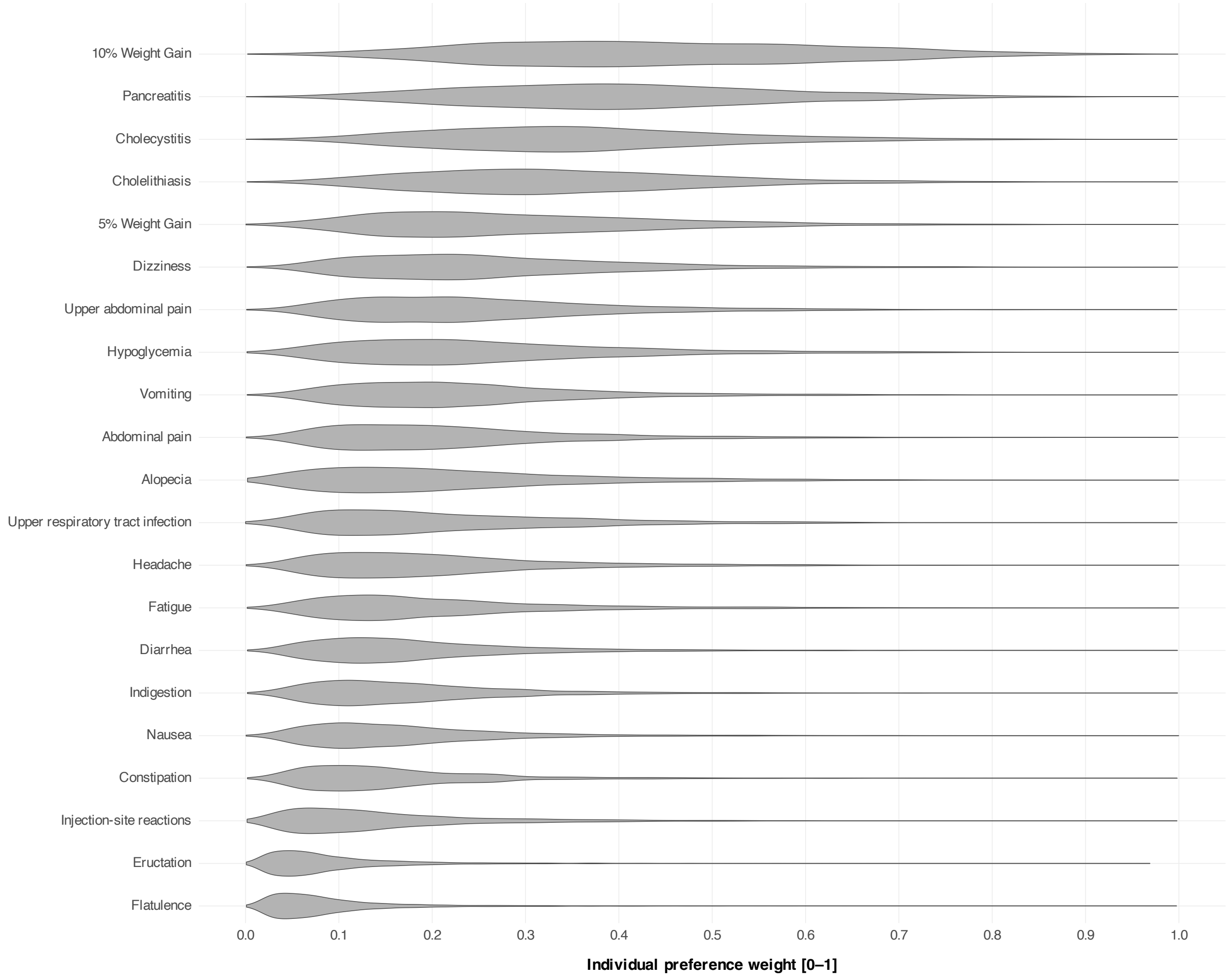

Violin plot showing the distribution of individual-level preference weights on a scale from 0 (least concerning) to 1 (most concerning). The width of each violin is proportional to the density of responses, with wider sections indicating a higher concentration of individuals assigning similar weights. Outcomes are ordered by the median preference weight.
